# Alpha coherence is a network signature of cognitive recovery from disorders of consciousness

**DOI:** 10.1101/2024.10.08.24314953

**Authors:** David W Zhou, Mary M Conte, William H Curley, Camille A Spencer-Salmon, Camille Chatelle, Eric S. Rosenthal, Yelena G Bodien, Jonathan D Victor, Nicholas D Schiff, Emery N Brown, Brian L Edlow

## Abstract

Alpha (8-12 Hz) frequency band oscillations are among the most informative features in electroencephalographic (EEG) assessment of patients with disorders of consciousness (DoC). Because interareal alpha synchrony is thought to facilitate long-range communication in healthy brains, coherence measures of resting-state alpha oscillations may provide insights into a patient’s capacity for higher-order cognition beyond channel-wise estimates of alpha power. In multi-channel EEG, global coherence methods may be used to augment standard spectral analysis methods by both estimating the strength and identifying the structure of coherent oscillatory networks. We performed global coherence analysis in 95 separate clinical EEG recordings (28 healthy controls and 33 patients with acute or chronic DoC, 25 of whom returned for follow-up) collected between two academic medical centers. We found that posterior alpha coherence is associated with recovery of higher-level cognition. We developed a measure of network organization, based on the distance between eigenvectors of the alpha cross-spectral matrix, that detects recovery of posterior alpha networks. In patients who have emerged from a minimally conscious state, we showed that coherence-based alpha networks are reconfigured prior to restoration of alpha power to resemble those seen in healthy controls. This alpha network measure performs well in classifying recovery from DoC (AUC = 0.78) compared to common representations of functional connectivity using the weighted phase lag index (AUC = 0.50 - 0.57). Lastly, we observed that activity within these alpha networks is suppressed during positive responses to task-based EEG command-following paradigms, supporting the potential utility of this biomarker to detect covert cognition. Our findings suggest that restored alpha networks may represent a sensitive early signature of cognitive recovery in patients with DoC. Therefore, network detection methods may augment the utility of EEG assessments for DoC.

## Introduction

Disorders of consciousness (DoC) result from disruptions to spatiotemporal oscillatory networks that underlie both arousal and impairment of processes supporting awareness and cognitive function. Bedside behavioral examination does not reliably provide an accurate diagnosis for some patient’s level of consciousness^1^. As a result, there is growing interest in the development of systems-specific indicators of the complex functions supporting arousal and awareness. Electroencephalographic (EEG) measures of the alpha frequency band (8-12 Hz) may be particularly well-suited to assess functions underlying awareness in patients with DoC because they are derived from activity implicated in conscious perception and cognition^2^. Unlike slow wave and delta (1-4 Hz) activity, which are considered indices of impaired arousal^3^, alpha is known to be generated by recurrent activity within thalamocortical circuits forming cortico-cortical networks within sensory systems^4,5^. Alpha activity reflects attentional selection and processing of sensory information^6^, and the phase cycles of alpha rhythms have been found to shape perceptual contents for downstream processing^7,8^. Posterior alpha connectivity has been linked to a variety of sensory and cognitive tasks including attentive and perceptual processes thought to be involved in the maintenance and function of conscious brain states^8,9^, as well as other aspects of cognition such as working memory and cognitive control^10–12^.

Previous work has shown that alpha-band measures are among the most important features in the classification of DoC, showing both prognostic utility^13,14^ and high performance in classification models compared to non-spectral measures such as evoked potentials, connectivity, and information theory^15^. While the functional importance of alpha-band activity is well-recognized, a main barrier to harnessing it for diagnosis, prognosis, and interpretation is the need to reduce its spatiotemporal complexity into a low-dimensional biomarker associated specifically with posterior alpha function^16^. Many correlational studies of the alpha rhythm in DoC have measured channel-based spectral power and some have measured coherence^17–19^, which typically involves dissecting network interactions into pairwise relationships between recording channels. Measures that quantify modes of neural activity in posterior alpha networks remain underexplored for DoC relative to sleep^20^ and anesthesia^21^. Here, we apply a dimensionality reduction technique called global coherence analysis to estimate coherence-based whole networks in multichannel EEG recordings^22–24^. Used in the clinical setting, posterior alpha measures may provide candidate biomarkers tailored to the assessment of brain systems supporting conscious awareness^16^.

In this dual-center, longitudinal study, we investigated the characteristics of EEG-derived alpha networks in patients with DoC at one or more time points after hospital admission. We explored two patient populations; one acute cohort in which patients with DoC were enrolled while in the intensive care unit (i.e., recordings obtained within first 2.5 weeks of admission) and subsequently followed during the chronic phase (5+ months post-injury), and a second cohort in which all recordings were obtained during the chronic stage. EEG recordings were obtained during resting-state periods and task-based paradigms. Both cohorts were recorded using

clinical-grade 10-20 EEG systems. The main objective of our study was to demonstrate the utility of global coherence analyses in addition to standard EEG-based spectral analyses methods for determination of potential recovery from DoC. A secondary objective was to study the network characteristics of alpha oscillations observed during the course of cognitive recovery.

Using multitaper spectral analysis, we investigated the frequency-domain and topographical profiles of EEG activity across patients in our study. Then, we applied global coherence analysis to measure the strength and organization of alpha networks. From global coherence outputs, we reconstructed spatial maps of regions contributing to the strongest coherence-based networks.

Next, we measured the similarity between alpha network configurations observed in patients throughout the course of recovery. We compared the diagnostic utility of network measures derived from global coherence with other types of network metrics, including those based on the weighted phase lag index (wPLI). Lastly, we provide proof-of-principle evidence that an alpha network-based signature of EEG command-following may be used to detect covert cognition in DoC patients.

## Materials and methods

We analyzed EEG recordings obtained in two previous studies from 33 patients and 28 healthy controls at two academic medical centers (site A: 19 patients, 16 healthy controls; site B: 14 patients, 12 healthy controls). The patient recruitment procedures, inclusion/exclusion criteria, EEG data acquisition procedures and study paradigms utilized in the two studies have been previously described^25,26^, and key methodological details are reiterated here.

### Subject recruitment and assessment

We enrolled 28 healthy controls without neurological, psychiatric, or medical disorders and 33 patients with DoC at two study sites (designated as sites A and B). The median patient age at time of injury was 27 (IQR = [23.5, 37.5]), and 22 patients were males. At site A, 19 patients were enrolled after being admitted to the intensive care unit (ICU) for acute severe traumatic brain injury (TBI). Of these, 15 patients were enrolled prospectively in the acute phase, 12 of whom received follow-ups during the chronic recovery phase ranging from 5 to 39 months post-injury. Four additional patients were enrolled at the follow-up stage and their acute clinical EEG and behavioral assessment data were obtained retrospectively. Criteria for inclusion at this site were: 1) age between 18 and 65 years; 2) head trauma with Glasgow Coma Scale score of 3-8; and 3) no eye opening for at least 24 hours. Criteria for exclusion were: 1) life expectancy less than 6 months as estimated by the treating physician; 2) prior neurodegenerative disease or severe brain injury; 3) body metal precluding MRI; and 4) lack of English fluency pre-injury.

At site B, 14 patients were admitted during the chronic recovery phase, 13 of whom had one or more follow-up study visits from 0.5 to 4.3 years (median: 2.1 years, IQR = [1.1, 3.1]) following the first visit. Including first study visits, EEG sessions at site B were conducted a median of 4.5 years (IQR = [2.1, 6.9]) after day of injury. Here, criteria for inclusion were: 1) age between 18 and 75 years; 2) the patient’s legal proxy’s fluency in English; 3) diagnosed with a severe, non-progressive brain injury; 4) medically stable; and 5) English fluency pre-injury.

Criteria for exclusion were: 1) a terminal illness diagnoses; 2) a diagnosis of refractory generalized epilepsy; 3) dependence on ventilator or dialysis; 4) evidence of Alzheimer’s disease or dementia pre-injury; 5) premorbid neuropsychiatric history; 6) significant acute or chronic illness; 7) participation in any investigational trial < 30 days prior to enrollment; and 8) requirement of physical restraints. All but three of patients in this cohort experienced a TBI and the remaining experienced either a hypoxic or hypoxic-ischemic brain injury.

At both study sites, behavioral assessments were performed with the Coma Recovery Scale-Revised (CRS-R)^27^ by one of the investigators (BLE, NDS). At site A, CRS-R assessments were performed immediately prior to each EEG recording session; at site B, they were made within 24 hours of the EEG recording session during the same visit. Patients who emerged from minimally conscious state (MCS) were assessed with the Confusion Assessment Protocol (CAP)^28^ to determine their confusion status (CS for confusional state or R-CS for recovered from confusional state). At site B, confusion status in patients who emerged from MCS was assessed with multiple instruments, including the CAP, Galveston Orientation and Amnesia Test (GOAT)^29^, and Mississippi Aphasia Screening Test (MAST)^30^. Using all test results, we labelled each patient’s status on the EEG recording day with one of seven diagnostic classes^31^: coma, unresponsive wakefulness syndrome/vegetative state (VS), MCS without language function (MCS-), MCS with language function (MCS+), CS, and R-CS. Diagnoses of coma, VS, MCS-, and MCS+ were derived from the CRS-R assessment, while diagnoses of CS and R-CS were derived from the CAP scores in patients who were deemed having emerged from MCS on the CRS-R. In some patients where full CAP results were not available (affecting n = 5 recordings), GOAT and MAST tests were used to differentiate CS from R-CS. Patients who did not receive sufficient additional GOAT or MAST testing to determine R-CS were classified as CS (See Table 1).

**Table 1.** Patient demographics and clinical information. Sessions within a single visit are separated by commas; sessions from multiple visits are separated by semicolons. Level of Consciousness (LoC) marked with * represents insufficient testing to ascertain recovery from confusional state (CS). LoC marked with ** determined by clinical emergence from CS. Initial visits marked with + represent earlier EEG sessions obtained from patients enrolled at follow-up. Days post-injury of EEG summary statistics were taken with the first date per visit (consecutive days) for each subject, with subject B20 received two follow-up visits. Other abbreviations: traumatic brain injury (TBI), vegetative state (VS), minimally conscious state (MCS), recovered from confusional state (R-CS).

| Subjects | Age Range at Injury | Sex | Etiology | Day(s) Post-Injury of EEG |  | LoC(s) at EEG |  | Total CRS-R Score(s) at EEG |  |
| --- | --- | --- | --- | --- | --- | --- | --- | --- | --- |
|  |  |  |  | Initial | Follow-up(s) | Initial | Follow-up(s) | Initial | Follow-up(s) |
| A1 | 26-30 | m | TBI | 17 | 206 | CS | R-CS | 23 | 23 |
| A2 | 21-25 | f | TBI | 4<sup>+</sup> | 224 | Coma | MCS- | 1 | 9 |
| A3 | 21-25 | m | TBI | 2 | 174 | MCS- | R-CS | 4 | 23 |
| A4 | 16-20 | f | TBI | 4 | 371 | Coma | R-CS | 1 | 23 |
| A5 | 16-20 | m | TBI | 10 | 576 | CS | R-CS | 22 | 23 |
| A6 | 31-35 | m | TBI | 16 | - | VS | - | 3 | - |
| A7 | 26-30 | f | TBI | 10 | 656 | MCS+ | R-CS | 11 | 23 |
| A8 | 41-45 | m | TBI | 14 | - | MCS+ | - | 18 | - |
| A9 | 31-35 | m | TBI | 8 | - | CS | - | 20 | - |
| A10 | 31-35 | m | TBI | 13 | - | MCS+ | - | 15 | - |
| A11 | 21-25 | m | TBI | 13 | - | MCS- | - | 10 | - |
| A12 | 21-25 | f | TBI | 14 | 187 | CS | R-CS | 22 | 23 |
| A13 | 16-20 | m | TBI | 6 | - | CS | - | 21 | - |
| A14 | 26-30 | m | TBI | 8 | 235 | MCS+ | R-CS | 7 | 23 |
| A15 | 31-35 | m | TBI | 4 | 191 | CS | R-CS | 14 | 23 |
| A16 | 26-30 | m | TBI | 5<sup>+</sup> | 182 | Coma | MCS+ | 1 | 15 |
| A17 | 21-25 | f | TBI | 4<sup>+</sup> | 160 | Coma | VS | 1 | 5 |
| A18 | 26-30 | m | TBI | 14<sup>+</sup> | 1172 | Coma | MCS- | 1 | 5 |
| A19 | 26-30 | m | TBI | 28 | - | CS | - | 18 | - |
| B20 | 56-60 | f | hypoxia | 377 | 602; 975 | MCS- | MCS+;<br>CS | 14 | 19; 22 |
| B21 | 21-25 | m | TBI | 753, 754 | 2316 | CS* | R-CS<br>** | 16 | 23 |
| B22 | 16-20 | m | TBI | 2915, 2916, 2917 | 3364 | MCS- | VS | 8 | 7 |
| B23 | 51-55 | m | TBI | 302 | 1434, 1435 | CS | MCS+ | 22 | 21 |
| B24 | 16-20 | f | TBI | 2412, 2413 | - | VS | - | 7 | - |
| B25 | 21-25 | m | TBI | 1976, 1977 | 2334, 2335 | CS* | MCS+ | 17 | 14 |
| B26 | 11-15 | f | TBI | 3858 | 4936 | MCS- | MCS- | 12 | 12 |
| B27 | 16-20 | m | TBI | 1590 | 2834 | MCS+ | MCS+ | 5 | 5 |
| B28 | 16-20 | f | TBI | 515 | 1669 | MCS- | CS* | 10 | 22 |
| B29 | 21-25 | m | TBI | 1085 | 1940 | MCS- | MCS- | 11 | 11 |
| B30 | 21-25 | m | TBI | 349 | 1156 | MCS+ | MCS+ | 17 | 11 |
| B31 | 51-55 | f | anoxia | 218 | 953 | CS | R-CS | 22 | 23 |
| B32 | 56-60 | m | anoxia | 679, 680 | 1435 | CS | CS | 23 | 23 |
| B33 | 21-25 | f | TBI | 5879 | 6045 | MCS- | VS | 10 | 3 |
| Summary (Median; IQR) | 24; 12 | - | - | 16; 671 | 964; 1645 | - | - | 12; 12 | 21; 13 |

All studies were approved by the Institutional Review Boards at the respective study sites. Written informed consent was obtained from healthy participants and surrogate decision-makers. Patients who recovered consciousness at follow-up provided informed consent to continue participating in the study.

### Study design

The two study sites used EEG protocols that incorporated a combination of resting-state and task-based paradigms^25,26,32^, but these protocols differed in detail. At site A, resting-state EEG epochs were selected from research recordings in prospectively enrolled patients and healthy controls. In a subset of patients, additional resting-state epochs were retrospectively obtained from clinical recordings. For the prospectively enrolled subjects, the experimental protocol consisted of five, five-minute resting-state epochs and four stimulus-based paradigms (including a CRS-R arousal protocol) administered in alternating fashion, with completion of the session contingent on clinical care demands of the ICU setting. All subjects were instructed to keep their eyes closed. During acute-stage EEGs, sedative, anxiolytic, and/or analgesic medications were administered to a subset of patients prior to or during EEG sessions at the discretion of the treating clinicians to ensure patient safety or comfort (See Supplementary Table 1.) At site B, resting-state and task-based EEG were acquired during study visits lasting several days in length. Resting-state data were extracted as either minutes-long epochs or as equal-length non-overlapping segments of at least three seconds uncontaminated by noise. Epoch and segment selections were made by a trained clinical electrophysiologist (MMC). Recordings at site A were not screened to exclude sleep, but when multiple clean resting-state blocks existed from a single session, the latest block of the session was used to maximize the likelihood of arousal from the CRS-R arousal protocol. At site B, the clinical neurophysiologist determined a patient’s level of alertness prior to initiating any task-based paradigms, and data selections were made in the absence of EEG sleep features. Together, all resting-state data were grouped into distinct recording sessions, totaling 67 sessions across 33 patients across both sites, 25 of whom received follow-up recordings (See Table 1). EEG recordings of site A healthy controls were made in the resting eyes-closed state, whereas site B controls were instructed to keep eyes open during resting and task-based paradigms.

The structure of the task- and stimulus-based paradigms differed between the two study sites. At site A, two stimulus-based paradigms comprising music and language were used. One task-based paradigm was also used, with an instruction to imagine opening/closing the right hand. These stimulus- and task-based paradigms consisted of six alternating 24-second rest and stimulus/task blocks. Within each block, subjects were instructed to perform the motor imagery task in response to four auditory stimuli delivered in succession, interspersed with 3-second periods to respond. At site B, patients were administered a battery of task-based paradigms, including instruction to imagine swimming, imagine playing tennis, and imagine opening/closing the left and/or right hand^33^. These paradigms were administered in sixteen alternating ‘on’ and ‘off’ commands, each of which preceded a silent response period of at least 10 seconds in duration. Stimuli and commands were delivered and synced to the EEG recording at both study sites.

### Data acquisition and pre-processing

EEG recordings at both study sites were digitized and amplified using Natus XLTEK EEG acquisition systems (San Carlos, CA), but different initial electrode configurations were used. At site A, all data were recorded with 19 electrodes arranged in the standard International 10-20 System montage (double-banana) with a 200, 250, or 256 Hz sampling frequency. At site B, the EEG was recorded using the conventional 19-channel montage as site A but augmented with additional leads (37 total). The EEG was recorded at sampling frequencies of 200, 250, or 256 Hz. To enable cross-site comparisons, the standard 19 channels that both datasets shared were isolated, reordered into a common sequence, and processed apart from the full channel set in a separate dataset. To harness data with higher spatial resolution for task-based analyses, we maintained a separate version of the original 37-channel dataset to run in the command-following analysis pipeline. At both study sites, the FCz electrode was used as the initial common reference. After a common channel set was selected, individual channels were band-passed filtered between 1 and 50 Hz using a third-order Butterworth, zero-phase shift filter. To improve measurement from local sources and remove contaminating activity at the reference electrode, all channels used in coherence analysis were re-referenced using a Laplacian montage constrained to provide each channel with between three and five nearest neighbors before being detrended in each epoch or segment. Data used in wPLI calculations were re-referenced using a common average reference, similar to other studies^34^. Prior to spectral and cross-spectral analyses, epochs were sectioned into consecutive windows of 3 seconds and selected segments were truncated to 3 seconds. (See Supplementary Figure 1 for schematic diagram of pre-processing steps.) All data were inspected visually in the frequency domain using channel-wise median spectrograms, and windows containing broadband electromyogenic or interference artifacts were rejected manually by a single investigator blinded to clinical and behavioral data (DWZ). In total, 6.4% of data were rejected. The median total data length used from individual recording sessions was 285 seconds (IQR = [123, 303]). Data were pre-processed and cleaned using custom code and the Chronux toolbox^35^ in MATLAB (Mathworks, Natick, MA).

## Data analysis and statistics

### Spectral, coherence, and cross-spectral analyses

Multitaper spectrograms and coherence matrices were calculated using the Chronux toolbox^35,36^ for 3-second non-overlapping windows. We used five tapers, 2 Hz frequency resolution, and 1/3 Hz grid spacing.

To identify coherence-based networks within the 19-channel multivariate time series, we estimated the cross-spectral matrix via the multitaper method and then applied principal components analysis, a procedure known as global coherence analysis^22,23^. We also used extended linear algebraic procedures described in ^24^ for computing frequency-domain bootstraps of the cross-spectral matrix and reconstruct coherent power spectral densities from principal components of the cross-spectra, portions of which are summarized here. We used the same multitaper parameters detailed above to compute Fourier coefficients 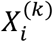of channel *i* ∈ *N* at frequency *f* for each non-overlapping time segment and *k* Slepian tapers. From the coefficients, we computed the *N* x *N* cross-spectral matrix 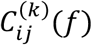 between the *i*-th and *j*-th channels for each respective taper and time segment:

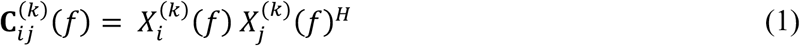

where 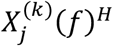 is the complex conjugate transpose of the vector of Fourier estimates 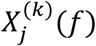

To make statistical inferences regarding network modes identified by global coherence, we applied a nonparametric resampling approach to the cross-spectral matrix. As detailed in^37^ and summarized here, this procedure generated surrogate cross-spectra using a non-overlapping block bootstrap in the frequency domain^38^. We generate *B* = 200 bootstrap replicates 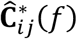 at each given frequency *f* by independently sampling from 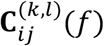 across *K* tapers and *L* time segments, then taking the mean across tapers and median across time segments for the real and imaginary components separately^39^. We computed eigendecompositions of surrogate cross-spectral matrices per frequency, per session.

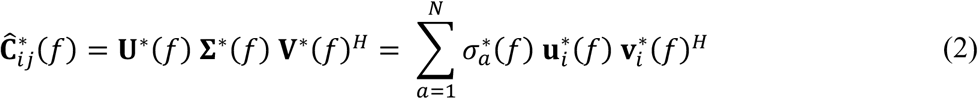

These bootstrapped principal eigenvalues and eigenvectors were used to obtain empirical distributions of global coherence (defined in ^22^ as *σ*_1_/ Σ^*N*^ *σ*_*a*_, the fraction of power explained by the principal eigenvalue) and the coherent power spectral density (defined in ^24^ as the channel-wise power density 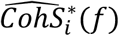 from the diagonal elements of 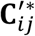, as well as the other downstream quantities defined below.

### Coherent subspace analysis

Because eigenvectors of the cross-spectral matrix encode channel-specific phase relationships, coherent activity of coordinated alpha generators can be thought to lie within a subspace defined by one or more principal eigenvectors^40^. By using identical channel sets in our clinical recordings, we sought to compare coherent subspaces using the principal angle

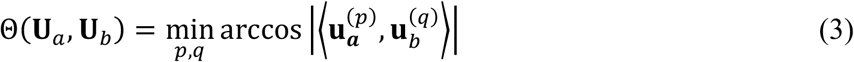

where 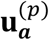 and 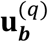 are the unitary *p*-th and *q*-th dimensional vectors within subspaces **U**_*a*_ and **U**_*b*_, respectively. Because vector dimensions are interchangeable, the distance measure does not take into account spatial organization of the channels; however, the relative positions of the recording sites influence the spatial specificity of the orthogonal eigenvectors and, therefore, distances between them. In this study we define principal subspaces (or “coherent subspaces”) using the single eigenvector representing the largest principal component. For each recording, we bootstrapped 200 subspace replicates 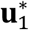 per EEG recording. Using these replicates, the bootstrap median angle between principal subspaces of separate recordings were computed as measures of distance between recordings (i.e. in separate subjects or sessions). Likewise, empirical median between subspace replicates of the same recording were computed to estimate variability of subspaces within the cross-spectral matrix.

### Additional distance measures and classification

To assess and compare the performance of other network measures as classifiers of disorders of consciousness, we computed three additional network representations, including two based on the wPLI. The first was a summary of channel-wise spectral power in vector form. The second and third were derived from pairwise wPLI defined by Vinck and colleagues^41^, which we computed from the bootstrapped cross-spectral estimates at the 10 Hz frequency that we used in global coherence analysis. To summarize wPLI networks in vector form, we computed the mean channel-wise wPLI (similar to Duclos and colleagues^34^) as well as the principal eigenvector of the wPLI pairwise matrix. As with coherent subspaces, distances between principal wPLI eigenvectors can be defined as the subspace angle or principal angle. For channel-wise power and channel-wise mean wPLI, we use cosine similarity for distance measurements. For all four distance measures, we assessed the effect sizes between within-group distances among controls on one hand and between-group distances from controls and patients of each DoC class on the other using Cohen’s *d*.

These four distance measures were used as scores for two types of classification: 1) patients with evidence of substantial cognitive recovery, defined as having CRS-R total score above or equal to 22 out of 23, and 2) patients who demonstrated R-CS. Receiver operating characteristic (ROC) curves and their AUC values were computed using each of the four distance measures in each of the two classification types.

### Group-level analysis

To estimate group-level empirical distributions of quantities generated during spectral and cross-spectral analysis, we used a hierarchical bootstrap approach. The first level bootstrap-resampled from patients. The second level bootstrap-resampled from time windows (in the case of spectral densities) or across replicates (in the case of cross-spectra) ^42,43^.

### Network-weighted spectral analysis

To represent the time-frequency activity of channels within a coherent subspace, we computed channel weights 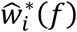 from estimates of the coherent power spectral density 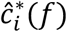.

We used these weights to define the network-weighted spectrum 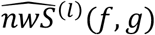, a channel-wise weighted average of multitaper spectra as a function of frequencies *g*.To summarize spectral activity weighted by channels contributing to a coherent basis, we computed channel weights 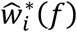 from estimates of the coherent power spectral density 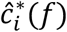. We use these weights to define the network-weighted spectrum 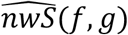, a channel-wise weighted average of multitaper spectra across frequencies *g*^24^.

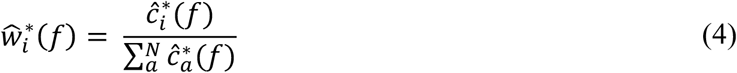

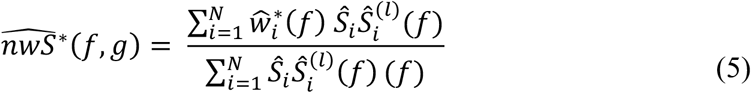

where *Ŝ*_*i*_(*f*)is the power spectrum at frequency *f*.

To estimate network-weighted alpha task suppression, empirical distributions of channel weights were computed at *f* = 10 Hz across both task ‘on’ and task ‘off’ data segments. Then, network-weighted spectra 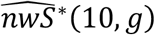 were estimated for each task ‘on’ and task ‘off’ window. Finally, bootstrap differences between task ‘on’ and task ‘off’ network-weighted spectral power were computed by subtracting activity between neighboring ‘on’ and ‘off’ periods, and then sampling across trials.

## Results

### Patient demographics and clinical characteristics

Our dataset consisted of 95 separate recording sessions from 61 subjects (28 healthy controls and 33 patients). 20 patients received two recordings each (the median). 25 of 33 patients returned for follow-ups between 5 months and 3 years after their first visit (median = 1.1, IQR = [0.5, 2.3] years). The patient cohort included 19 acute individuals at site A and 14 chronic individuals at site B. The median age across both sites was 24 (IQR: 12 years). 11 patients were female (five and six at sites A and B, respectively). Patients A1, A3-15, and A19 at site A were prospectively enrolled in the acute setting as part of a previous study^25^, and the remaining four site A patients were enrolled at follow-up. All site B patients were enrolled in the chronic setting. Altogether, 25 patients were studied at follow-up, and the rest did not complete follow-up for logistical or medical reasons, or due to being deceased. Acute EEG recordings at site A were taken median 10 days post-injury (IQR: 9.5 days), and the first chronic EEG recordings at site B were taken median 919 days post-injury (IQR: 1891.5 days). Follow-up EEG recordings at site A were taken median 215 days post-injury (IQR: 236.5 days), and those at site B were taken 1804.5 days post-injury (IQR: 1483.5 days). Supplementary Table 1 lists sedative, anxiolytic, and analgesic medications administered before and/or during acute EEG sessions at study site A. Three site A patients received continuous sedative infusions before or during EEG their recordings, and seven site A patients received intravenous analgesic or anxiolytic boluses.

Of the 95 total EEG sessions, 5 occurred while patients were in coma, 6 while VS, 14 while MCS- and MCS+ respectively, 18 while CS, and 10 while R-CS. At their first EEG, patients spanned the following diagnostic classes: coma (*n* = 5), VS (*n* = 2), MCS- (*n* = 8), MCS+ (*n* = 6), and CS (*n* = 12); at follow-up, the patients remaining in the study spanned the following diagnostic classes: VS (*n* = 3), MCS- (*n* = 4), MCS+ (*n* = 6), CS (*n* = 3), and R-CS (*n* = 10). (Table 1). CRS-R assessments spanned the entire range of possible outcomes with scores ranging from 1 to 23 (Fig. 3A). The upper end of this range included both CS and R-CS patients. (One patient (B20) received two follow-up testing sessions and was included in two of the follow-up diagnostic classes.) At site A, four patients (A2, A16, A17, A18) were in comatose state. This was assessed by the following criteria: lack of eye opening, tracking and fixation when eyelids were manually opened, purposeful movement, or response to verbal or noxious stimulation rather than CRS-R^44^. At site B, four subject visits did not have sufficient testing to ascertain emergence from CS using the CAP, GOAT, or MAST although CRS-R testing indicated emergence from MCS. Of those, one (B21) was nevertheless diagnosed as R-CS at follow-up due to full behavioral orientation to space and time.

### Detection of oscillatory networks in clinical EEG using global coherence analysis

In contrast to standard spectral analysis, global coherence analysis uses a dimensionality reduction approach to isolate power belonging to a coherent basis within the multi-channel data (Fig. 1A). A single principal component of the alpha cross-spectral matrix may summarize more than half of the total power at a given frequency (Figs. 1D, F). Alpha spectral power canonically has a posterior topographical profile (Figs. 1C, 2A-C). By reconstructing the channel-wise estimates of coherent activity, global coherence analysis is able to produce a topographical profile of activity belonging to a network synchronized at a given frequency (Fig. 1H).

**Figure 1.**
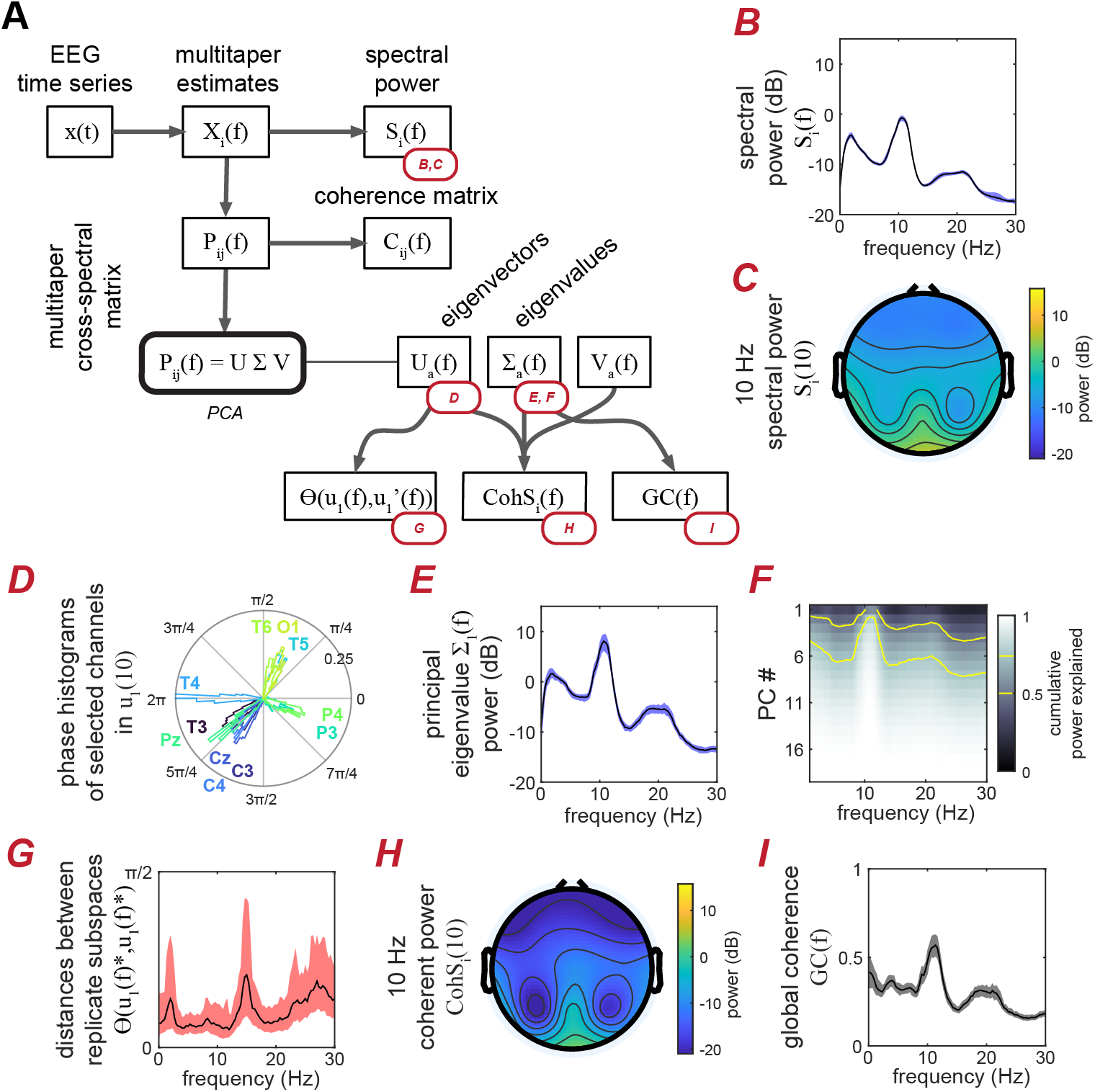
Using cross-spectral matrix eigendecomposition to extend standard spectral analytical methods. (A) The relationship between multitaper spectral analysis and global coherence analysis, with their respective outputs. Multitaper spectral analysis yields spectral and cross-spectral estimates, which can be plotted as spectra (channel-averaged here) or topographical plots (B-C). By performing principal component analysis on the cross-spectrum of a 19-channel EEG recording of a healthy control, the majority of total 10-Hz power can be summarized within the first principal component (D). The resulting eigenvalues and eigenvectors represent power of coherent bases (E) and phase relationships between channels in the network (I), respectively. These outputs can be used to produce additional quantities such as the global coherence (GC) (F), the coherent power (CohS) (G), and the angular distance (Θ) (H). Shaded boundaries represent the 5% and 95% confidence intervals of the median standard error.

We found that global coherence measures yielded higher signal-to-noise characteristics, producing larger alpha peaks relative to background activity for the canonical frequencies of interest. At 10 Hz, the principal, or largest, eigenvalues are up to 10 dB greater in power than channel mean estimates of spectral power, while widening the contrast between peak and background power by up to 5 dB (Figs. 1B, E). We also found that the network reconstruction of coherent power, an output of global coherence analysis, yields sharper contrast in its topography than standard 10-Hz spectral power (Fig. 1C, H). Across our dataset, we saw that coherent power aids in visualization of frequency bands such as alpha compared to classic spectral power (Figs. 3B, C), while global coherence lowers the background of potentially spurious coherence which often contaminates classic pairwise coherence measures (Figs. 3D, E). Comparing measures computed from recordings in healthy controls between the two experimental sites, we found no difference in the frequency-domain and topographical characteristics of alpha-band spectral, coherent power, or global coherence features (Supp. Fig. 2).

**Figure 2.**
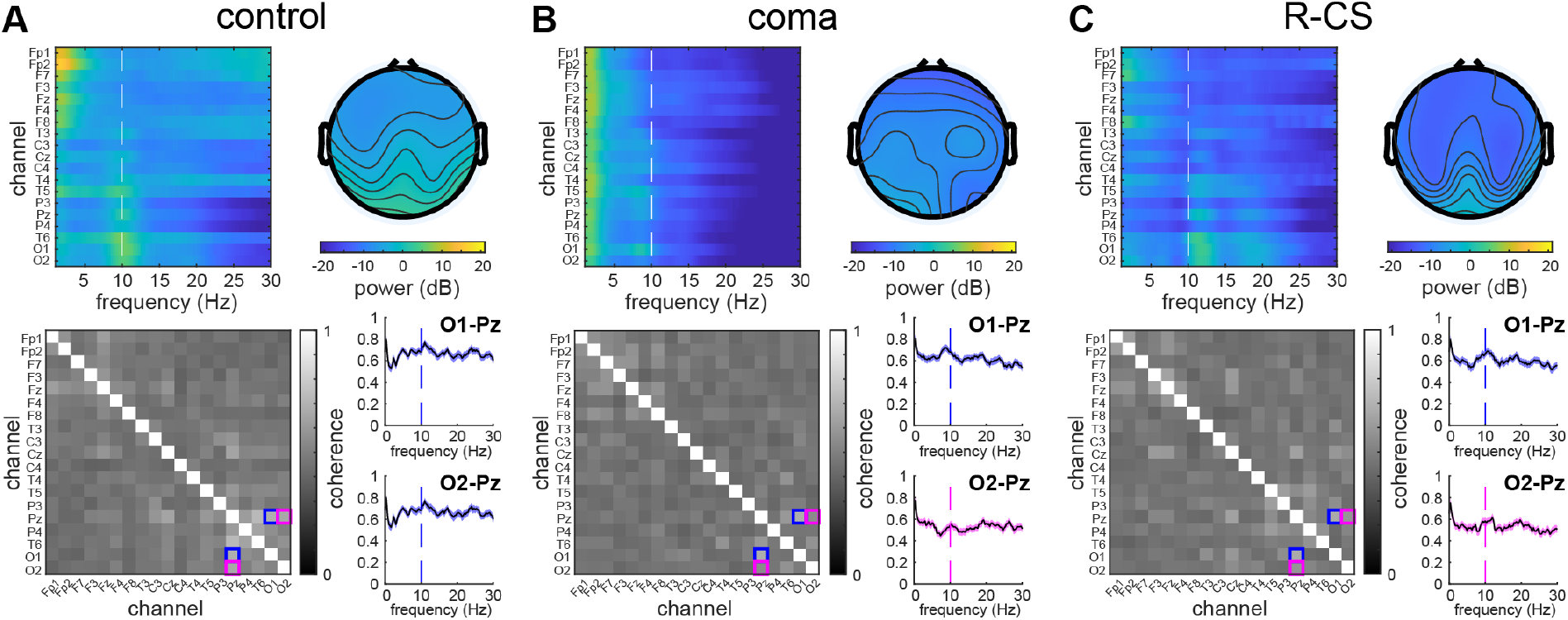
Spectral and coherence analysis of healthy subjects and patients with disorders of consciousness reveals posterior alpha component in conscious subjects. (top) Spectral topographical characteristics of 19-channel EEG recordings in a healthy control (A), an acutely comatose TBI patient (B), and a patient (C) after emerging from the confusional state. (bottom) Coherence matrices and coherence spectra between posterior channels recorded in the same subjects. Pairwise coherence plots and their corresponding matrix element are linked by blue and magenta colors.

**Figure 3.**
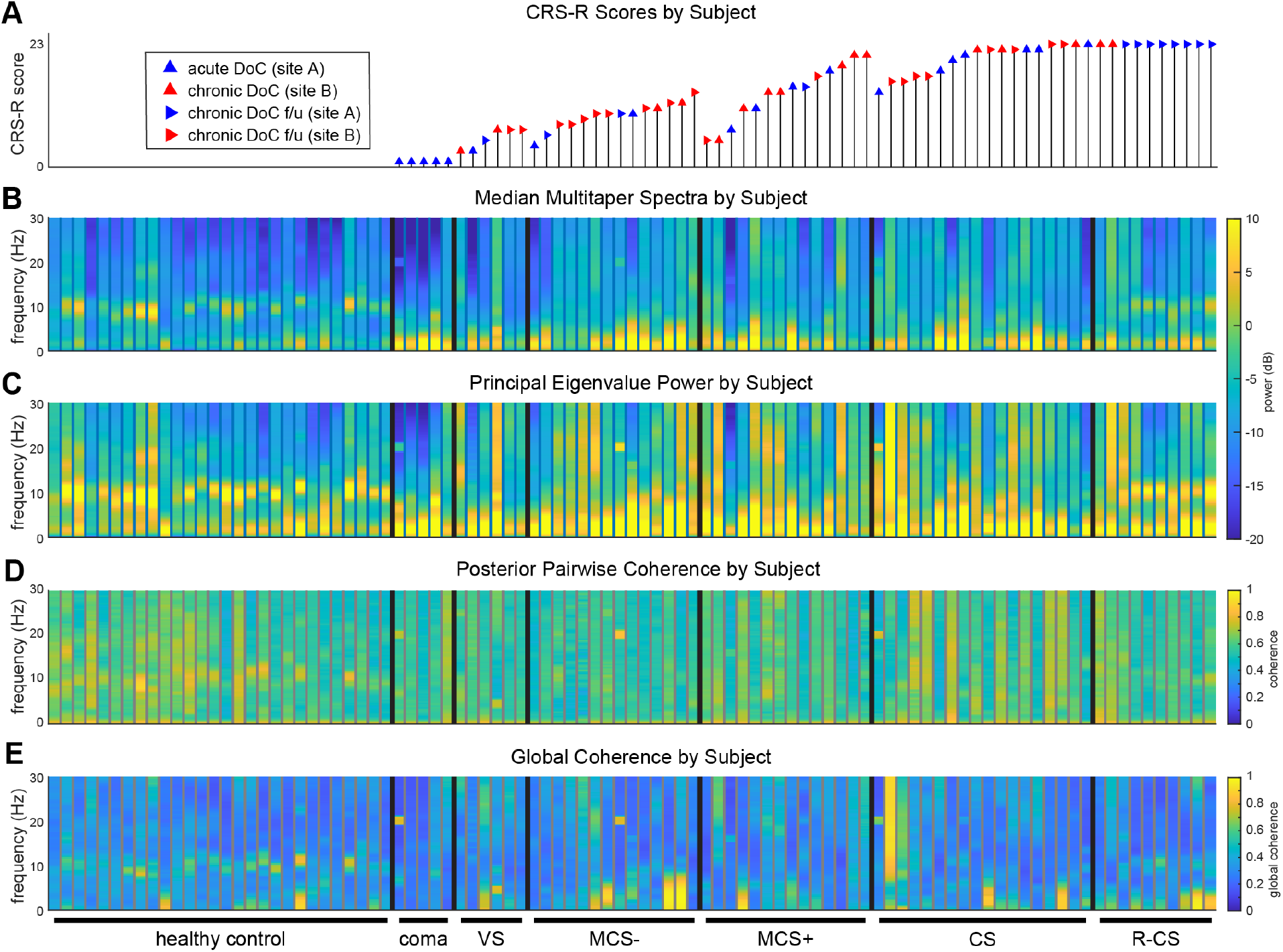
Subject-level comparisons between median multitaper spectra, the spectra of the principal eigenvalue, and the global coherence. (A) CRS-R total scores associated with each recording in the dataset. (B) The mean multitaper spectral power was computed across channels, and the median was computed across time. (C) Power of the principal eigenvalue. As in median spectral power, large eigenvalues in the alpha band can be found in most healthy controls and R-CS patients, but are lacking in patients in DoC groups between the coma to CS levels. (D) Mean pairwise coherence among the posterior O1-Pz and O2-Pz pairs. (E) Global coherence is defined as the proportion of the total power represented by the principal eigenvalue.

In addition to network strength, global coherence analysis robustly captures network structure in the form of motifs that we term “coherent subspaces”. In contrast to standard pairwise coherence measures that summarize synchronous relationships between channel pairs, global coherence analysis captures network synchrony within the entire channel set within the eigenvectors of the cross-spectral matrix (Fig. 1A). These eigenvectors, which define the coherent subspace, are complex-valued objects in which each element describes the phase of a constituent channel in relation to the network as a whole (Fig. 1I). By resampling the multitaper cross-spectral data, we consistently detect similar subspaces within cross-spectra belonging to frequency bands such as theta, alpha, and beta that are characterized by oscillations with strong cortical synchrony (Fig. 1H).

### Spectral and coherent dynamics across disorders of consciousness

We found that posterior alpha dynamics are a dominant feature of conscious brain activity in healthy controls and patients with R-CS. We observed that the alpha topography of healthy controls is a posteromedial phenomenon of the resting eyes-closed state (Fig. 2A). Alpha power in this posterior region is absent in DoC but present in R-CS (Figs. 2B, C and 4B). At the individual level, alpha spectral and coherent features were consistently present in healthy controls but generally absent in DoC (Fig. 3). Coherent power and global coherence measures revealed larger group-level differences between DoC patients and controls compared to spectral power and pairwise coherence measures, respectively. (Fig. 4A, top). In contrast, robust differences in spectral, coherent power, or global coherence between control and R-CS groups could not be identified in the alpha frequency band (Fig. 4A, bottom). Although the CS group can be differentiated from R-CS patients by lower spectral power in the alpha range (Fig. 3 and Supp. Fig. 5), a posterior topography in coherent power exists in both groups (Fig. 4B and Supp. Fig. 3). We also found that increases in posterior coherent power can be seen in individuals with increased level of consciousness at follow-up (Supp. Fig. 6). Patients comprising the CS and R-CS groups did not substantially differ in age (means of 33.9 and 28.8, standard deviations of 15.0 and 12.0, respectively).

**Figure 4.**
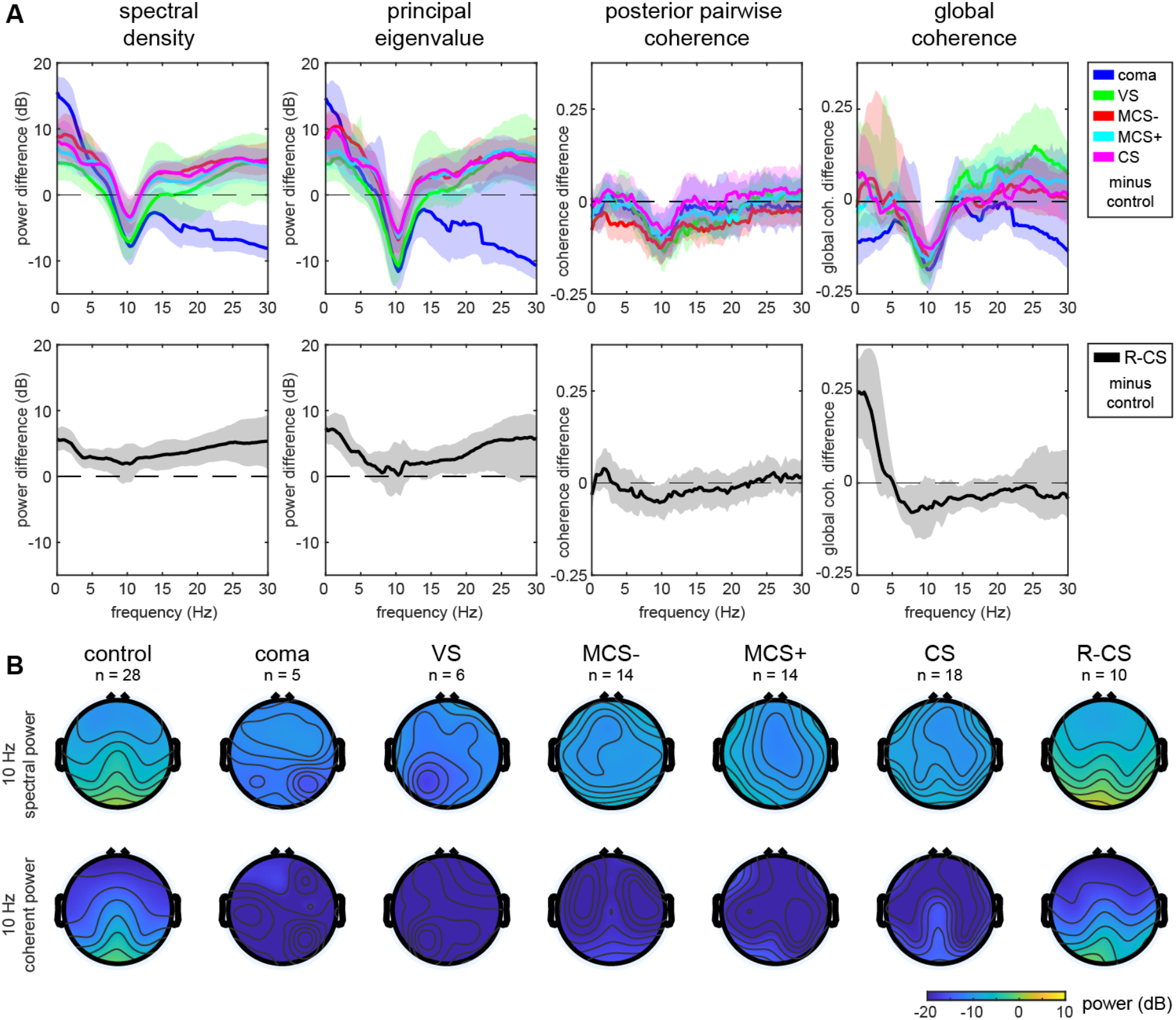
Spectral, cross-spectral, and global coherence features are sensitive to loss of posterior alpha activity in DoC. (A) Group-level differences between DoCs and controls (top) reveal loss of posterior alpha activity across measures. Shaded boundaries denote 5% and 95% confidence intervals. Group-level differences between R-CS and controls (bottom) do not exceed confidence bounds for spectral density, principal eigenvalue, or global coherence estimates. (B) Group-level median 10-Hz spectral (top) and coherent power (bottom) topographical plots across diagnostic classes, including coma, vegetative state (VS), +/-subclasses of minimally conscious states (MCS), confusional state (CS), and recovered from confusional state (R-CS). The posterior alpha component is visible in control, CS, and R-CS groups.

**Figure 5.**
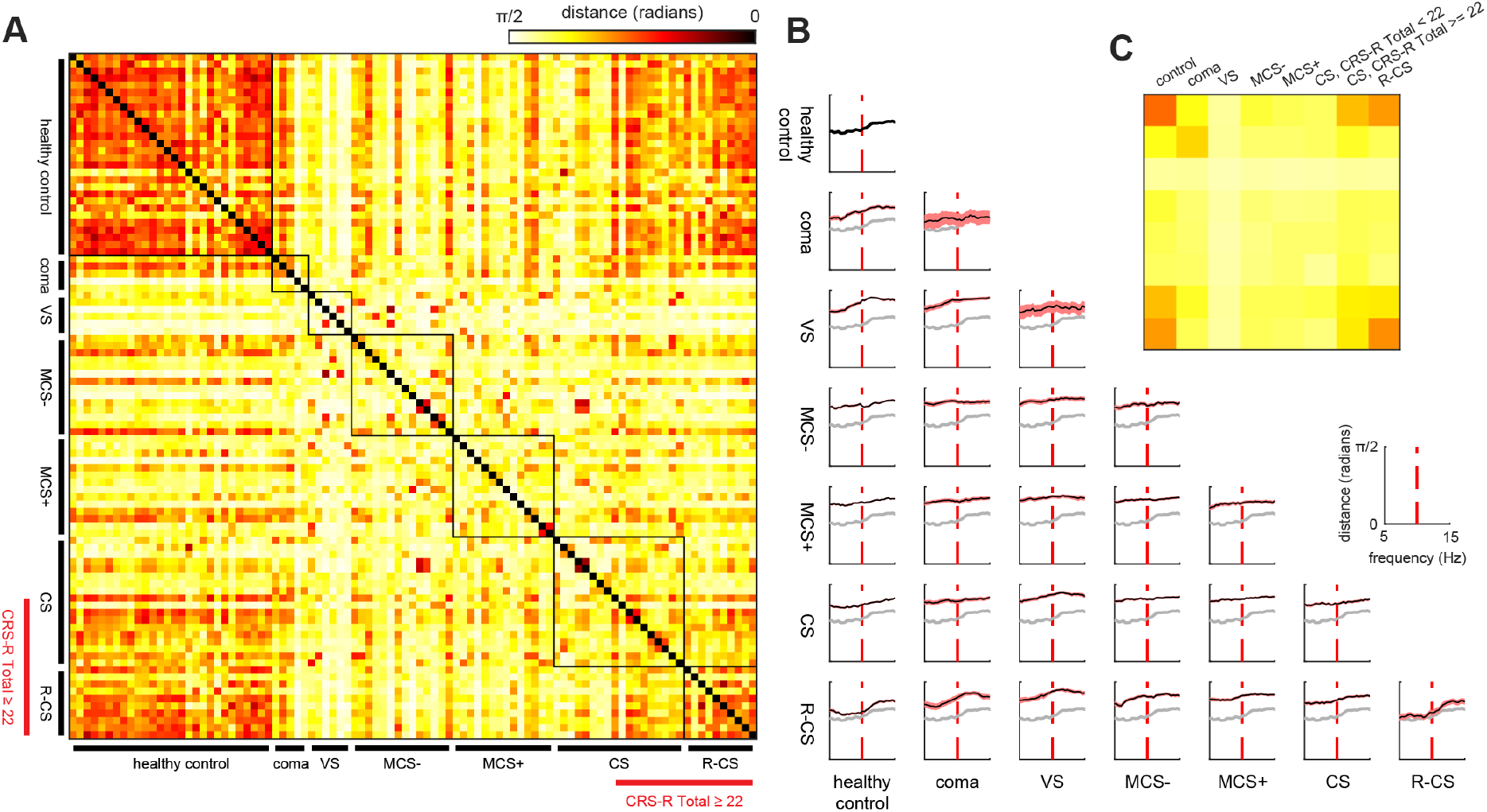
Between-subject and between-group distances measured using the principal angle between coherent subspaces. (A) The principal angle between principal eigenvectors (subspaces) from two subjects represents a distance measure ranging from 0 to pi/2 at the maximum. (B) Median distances across frequency between and within subspaces drawn from each subject group. In each plot, median distances between healthy control subspaces are plotted in gray. (C) At the 10-Hz frequency, distances are lowest between healthy controls and patients with high levels of recovery (both R-CS patients and CS patients with CRS-R total scores greater or equal to 22 during the visit). Consistent network structure representing healthy alpha network activity is represented by low within-group distances for controls and R-CS.

**Figure 6.**
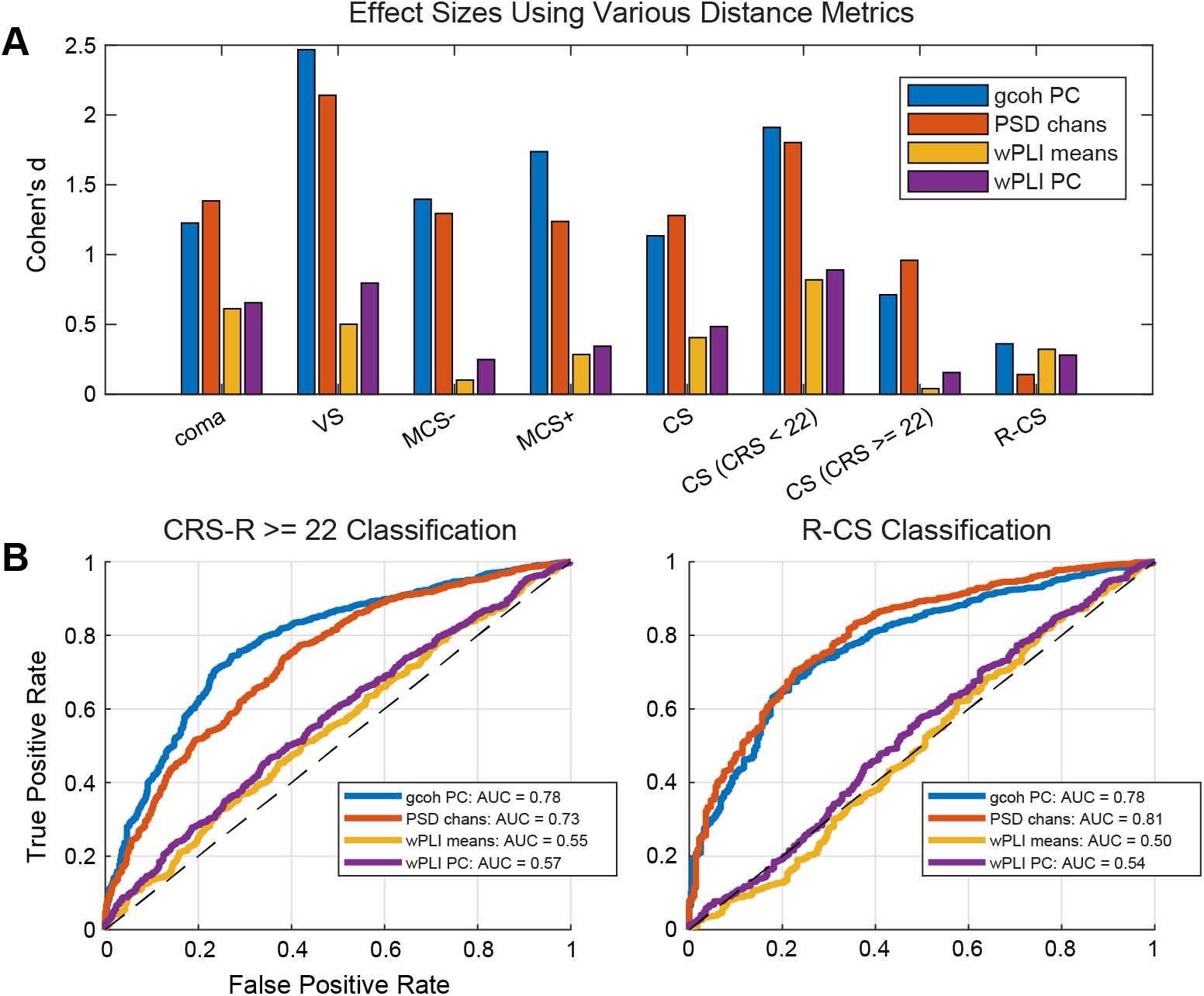
Coherent subspace and other distance measures in classification of recovery to high-functioning confusional state and beyond. (A) Effect sizes (Cohen’s *d*) comparing within-group distances among healthy controls and between-group distances between controls and DoC patient classes using four types of brain measures: 10-Hz coherent subspaces (gcoh PC), mean channel-wise 10-Hz multitaper power spectral density (PSD chans), mean channel-wise weighted phase lag index (wPLI means), and the principal eigenvector of the wPLI matrix (wPLI PC). Coherent subspaces and channel-wise alpha spectral power exhibit the largest effects in representing standardized differences between multichannel EEG of healthy and DoC populations. (B) Comparative performance of the multichannel measures as binary classifier scores for high CRS-R (scores greater than or equal to 22) or R-CS levels. Both coherent subspaces and channel-wise power effectively classify R-CS, but channel-wise power’s accuracy is lower for CRS-R >= 22 patients, evidenced by the AUC values.

Frequency-domain representations of these measures reveal clearer peak structure in the alpha band compared to theta and beta (Supp. Fig. 4). Spectral and coherent power estimates at theta and beta frequencies were not reliable measures of loss of consciousness in terms of statistically significant differences between DoC groups and both controls and R-CS (Supp. Fig. 5). In contrast, alpha coherent power was significantly lower in DoC patients than that of either controls and R-CS subjects, while spectral power and global coherence measurements in this band were significantly different only between DoC and R-CS or controls, respectively (Supp. Fig. 5). In the delta frequency band, spectral and coherent power were higher in DoC patients than in controls could not reliably distinguish between levels of consciousness (Supp. Figs. 4 and 5). Both spectral and coherent delta power have a fronto-medial topography but could not be associated with any specific levels of consciousness among patients (Supp. Fig. 3).

### Measuring alpha network similarity and classifying DoC recovery using coherent subspaces

We investigated how alpha network configurations varied across our patient population using our concept of the coherent subspace. Coherent subspaces of 10-Hz cross-spectral matrices from EEGs of healthy controls exhibited low within-group distances, indicating highly consistent network structure (Fig. 5). When compared to patient groups, the subspaces of healthy controls shared the greatest similarity with those of R-CS as well as a subset of CS patients who scored a CRS-R total of at least 22 (Fig. 5C). Within this subset of CS patients, we found a dissociation between alpha power and alpha network similarity to controls, where low alpha spectral power (Fig. 3B) seemed to coincide with alpha coherent subspaces with higher similarity to those of control subjects (Fig. 5C).

To evaluate the ability of alpha-band coherent subspace similarity to classify DoC, we compared its performance against other representations of network topography. Functional connectivity studies in states of consciousness commonly use real-valued pairwise measures such as the phase lag index and its variants, like wPLI^34,45,46^. To reduce the dimensionality and assess the participation of a spatial node to the rest of the brain network, these approaches average wPLI estimates between one region and all others. Alternatively, a PCA-based approach may be used to represent a network as a dominant basis within the wPLI pairwise matrix. Lastly, channel-wise spectral estimates also yield a vector-based readout of network topography without using phase information. In total, we compared four types of network representations in distance-based classifications of DoC: the coherent subspace, channel-wise spectral power, channel-wise mean wPLI, and the principal component of the pairwise wPLI matrix.

First, we assessed the ability of these measures to separate the diagnostic classes. Specifically, we compared between-group distances between healthy controls and various DoC classes, with the within-group distances. We found that channel-wise spectral power and coherent subspaces, when used to measure distance between subjects from different groups, demonstrate greatest separation (Fig. 6A). These sizable effects were specific to DoCs and were diminished in R-CS to levels close to those of wPLI-based measures. To evaluate the potential utility of different network measures to predict degree of recovery, we then used each patient’s distance from healthy controls as scores in binary classifiers of either patients with high CRS-R score (greater than or equal to 22) or R-CS patients (Fig. 6B). We found that both coherent subspaces and channel-wise power perform well as scores to classify R-CS, but coherent subspaces are more accurate when used to classify patients scoring highly on the CRS-R, as measured by the AUC. Consistent with the observation that emergence of alpha network similarity to healthy controls typically occurs earlier in recovery than increases in posterior alpha power, coherent alpha subspaces may have greater sensitivity to high CRS-R patients compared to channel-wise power. By contrast, we found that both versions of wPLI-based network representation exhibit lower classification accuracy than alternatives, similar to other studies of wPLI-based classification accuracy^34,46,47^.

We then compared the performance of alpha-band coherent subspaces in classifying patients exhibiting high levels of recovery against that of other canonical frequency bands such as theta and beta (Supp. Fig. 7). At these frequencies, the classification accuracies were 0.64 and 0.60, respectively, below that of alpha coherence subspaces at 0.78 (Fig. 6B). The subject-wise similarity matrix of theta subspaces reveals some similarity between healthy controls and R-CS, as well as clusters of similarity loosely distributed throughout the patient population. For beta, healthy controls and most R-CS patients were not similar in their subspace orientations, and the group-level similarity was weak compared to alpha (Supp. Fig. 7).

**Figure 7.**
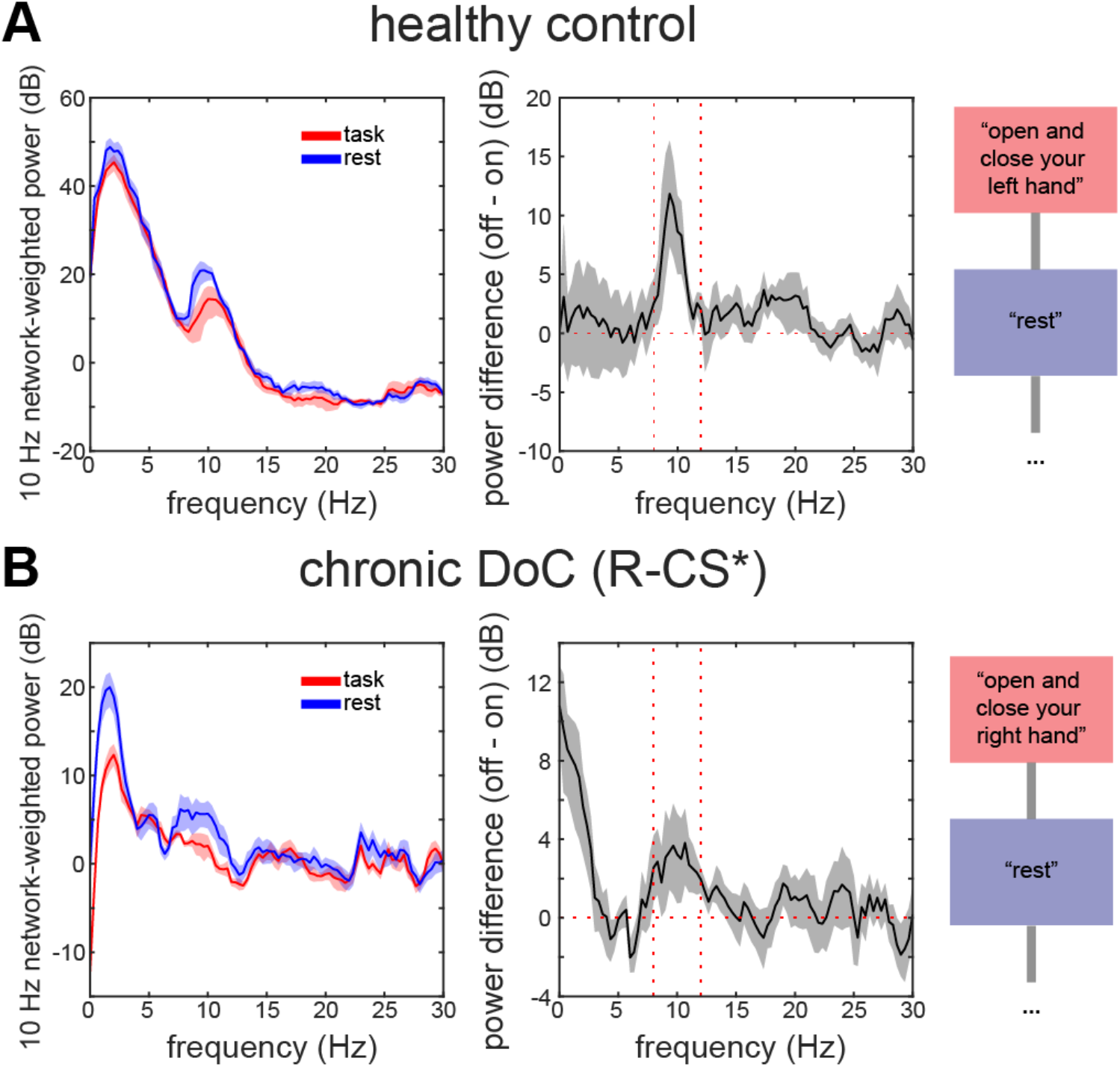
Examples of network-weighted alpha task suppression in a healthy control and a patient with chronic DoC. During the tasks shown, both subjects were instructed to open and close either the left or right hand, followed by instruction to rest (right). The spectral power weighted by channels representing global coherence networks (left) was suppressed in the alpha range (8-12 Hz) when subjects performed the task, as shown by bootstrap differences between network weighted power observed during neighboring tasks (middle). * denotes clinical emergence from CS but was not assessed with behavioral assessment.

### Alpha network activity yields evidence of command-following

Previous studies have shown that alpha oscillations are suppressed when conscious subjects silently follow commands^26,48^. Here, we extend the concept of alpha task suppression to task-relevant alpha networks by using channel weights identified by the coherent network reconstruction method. We used a command-following dataset obtained during the same visits from which the resting-state data at site B were collected. Thirteen of the 33 patients in this dataset were also reported in Curley et al^26^.

We found that alpha networks can be modulated by task performance in healthy controls and recovered patients. Figure 7 depicts task-related network dynamics in two subjects, a healthy control and a chronic-setting patient who exhibited sufficient behavioral markers to be labelled R-CS. All patients previously identified in the Curley et al. study as “command-following positive” also exhibited task modulation of the alpha network (Supp. Table 2). However, we also found that three of four patients previously reported with negative responses (B22, B27, and B28) scored positively in the present study. These four patients were all in MCS+/-states, with CRS-R total scores of 5, 8, 10, and 12. Four of 16 patients exhibited positive responses in more than one tested paradigm. Among those patients were two VS patients, one CS patient, and one R-CS patient who exhibited positive responses in the majority of task blocks tested. These patients had a total CRS-R scores of 7, 7, 22, and 23 respectively. In 15 healthy controls, 13 exhibited positive alpha network responses in at least two out of three task paradigms, with eight healthy controls being positive in all three. Importantly, the command-following results reported here required use of the original 37-channel set collected at the study site; the command-following signature did not retain sensitivity when data were reduced to the 19-channel montage of the above resting-state analyses.

## Discussion

In this multi-center study of EEG network dynamics in patients with DoC, we found that spatially coherent alpha activity is less prominent in DoC and restored in patients who recover consciousness. Using techniques developed from global coherence analysis, we measured whole networks in the alpha band across the spectrum of DoC. In contrast to loss of arousal, which is thought to coincide with increases in slow-delta activity during traumatic coma^49^, loss of higher-level cognitive function may be associated with attenuation of posterior alpha networks. Our findings show, in a subset of CS patients who score highly on the CRS-R, that coherent alpha patterns similar to those seen in healthy controls and R-CS patients begins to emerge in patients with DoC who recover higher-level cognitive function (i.e., CS and R-CS) before alpha power is recovered, suggesting that alpha synchrony may be an early signature of cognitive recovery. Finally, we found that the topographic regions that contribute to alpha global coherence exhibit task suppression, lending support to alpha network dynamics as a task-related phenomenon indicating cognition. Overall, our results demonstrate the diagnostic utility of global coherence analysis as sensitive, network-based descriptions of oscillatory dynamics compared to standard spectral and coherence measures.

### Global coherence analysis as a network neuroscience approach for the clinic

Global coherence analysis was originally posed as a multichannel coherence estimation technique^22^. In this study, we apply it more broadly as a decomposition technique for oscillatory network identification and comparison^24,50^. By orthogonalizing the cross-spectrum, it is possible to interpret principal components as a set of network-level physiological generators contributing, in the absence of common voltage sources, to the dominant synchronous activity in the channel set. On the other hand, activity represented by smaller principal components may contain either networked sources explaining lesser variance or noise that comprise the remaining activity. By analyzing the largest component, we rely on the assumption that the posterior alpha component typically contributes the greatest amount of coherent alpha activity in healthy waking brains, consistent with the vast literature on alpha rhythms since the inception of EEG^51^. Physiological impairment in the sources underlying this component would lead to orientation changes in the basis vector, attenuation of the global coherence magnitude, or both.

Although various functional connectivity methods have been used previously for classification of DoC^52–55^, there is not currently a consensus view favoring any network measure over others with respect to interpretational value or diagnostic accuracy. By preserving network representations as an integrated set of complex-valued phase relationships, our approach conceptually differs from common methods in functional connectivity such as wPLI, which produces separate pairwise calculations without retaining their covarying structure among channels. Rather than assigning strengths to individual network edges based on a coupling function, coherent subspaces represent estimates of all channels’ phase positions within a network represented by the principal component. Using the subspace framework, we were able to precisely estimate the structure of a network in reduced dimensionality and achieve larger statistical effects at the group level compared to measures based on wPLI.

### Potential mechanistic basis of reduced alpha coherence in DoC

We may interpret these findings on alpha functional networks in DoC in the context of sensory neurophysiology. Alpha rhythms are thought to be generated by recurrent thalamocortical interactions that create coherence through feedback from the cortex^56,57^. Thalamocortical circuitry constitutes the upper portion of the mesocircuit model, in which disruptions have been linked to DoC^58–60^. In this model, the central thalamus, which receives input from the ascending arousal network^61^, mediates broad arousal of the frontal cortex during the wake state^62^. Evidence in macaques has shown restoration of long-range fronto-parietal alpha coherence when the central thalamus is electrically stimulated at high frequency^63^. Our study places particular focus on a posterior region spanning parietal and occipital cortices as where coherent alpha networks associated with consciousness are situated. Although our recordings cannot spatially resolve cortical sources of posterior alpha, they point to a posterior and parietal spatial distribution situated in conscious subjects among electrodes O1, O2, and Pz of the 10-20 system (Fig. 1C). The mesocircuit, which comprises connections from anterior and intralaminar thalamus to anterior cortical regions to a greater degree, may not directly drive the posterior alpha network but instead interface with it indirectly through the frontoparietal network. Functional MRI studies have suggested that DoC may be associated with lower functional connectivity in posterior nodes of the frontoparietal or the default mode network, such as the precuneus or posterior cingulate^64,65^.

Brain injuries, particularly those of traumatic nature, may disrupt coordination between posterior thalamocortical regions through a number of mechanisms, including: 1) disruption of long-range feedback interactions from higher-order cortex via a frontoparietal network, possibly by thalamic means^11,65–67^, 2) disruption of short-range feedback by interrupting traveling alpha^68^, 3) structural or functional deafferentation of posterior cortex due to degradation of thalamic inputs^16,69,70^, or 4) dysregulation of ascending or descending arousal to sensory thalamic nuclei, disengaging the high-threshold bursting state^71^. These possibilities are not mutually exclusive. For example, deafferentation of thalamus may impair frontoparietal network feedback, because long-range coherence has been shown to involve thalamus^63,72^. Alternatively, focal injury to fibers between occipitoparietal cortex and thalamus may dysregulate alpha by degrading mutual excitation to both regions^16^. Separately, forms of coma where alpha activity is present, termed “alpha pattern coma” (APC) are known to occur^73^. APC, observed mostly in acute settings, is not universal to DoC and has been characterized notably by a widespread bilateral spatial distribution that distinguishes it from the dynamics reported here^74^. The circuit layout and mechanisms of APC are unclear. Other states of unconsciousness in which alpha is increased relative to baseline include propofol anesthesia, which has been shown to be associated with a separate set of frontal thalamocortical connections and, thus, linked to distinct mechanisms^24^.

### Partial restoration of posterior alpha networks

Coherent alpha networks may serve as a neural activity biomarker of cognitive recovery, augmenting diagnostic capability when neuropsychological assessments do not detect emergence from DoC. We found that alpha power and synchrony are dissociable in CS patients with high levels of recovery (with a CRS-R total of 22 or above), who may have been grouped into CS or R-CS classes. Before alpha rhythms regain full power in R-CS, activity in these high-scoring CS patients appear to synchronize and organize in the posterior region, resulting in orientations of the coherent subspace resembling the posterior alpha seen in healthy individuals. A study by Shah et al. identified a posterior medial complex in which high delta-alpha ratios (i.e., low relative alpha) are associated with severity of post-traumatic confusional state^16^, supporting our finding that alpha dysfunction within this region may cause patients in severe confusional states to exhibit disorganized alpha activity. While age may correlate with lower alpha power in the broader population, our CS and R-CS patients have nearly identical age distributions. The physiological basis of alpha network synchrony emerging in settings of low alpha-band power is unclear but may indicate partial dendritic reconnection between thalamus and cortex prior to full afferentation^75^. The neurophysiological mechanisms underlying alpha network changes merit further investigation in the context of cognitively impaired states.

### Comparison to theta and beta coherent network activity

In addition to alpha, theta and beta oscillations also form coherence-based networks generated by cortical-subcortical circuitry^76,77^. These three oscillatory bands are major components of the ‘ABCD’ model of neurological recovery from DoC, whereby the course of recovery follows an ascending hierarchy of oscillatory states, with theta preceding alpha and beta^44,48^. In this model, alpha oscillations appear only in ‘D’-type dynamics, occurring closest to the recovery stage. A previous analysis of a subgroup of patients included in the present study showed correspondence between ABCD classifications and oscillatory dynamics in these frequency bands^44^. Our findings further demonstrate that, in addition to alpha oscillations, theta and beta oscillations underlying ABCD dynamics may have coherent network representations as well. Some degree of network similarity is recovered in theta and beta frequency subspaces across DoC at the group-level (Fig. 5B and Supp. Fig. 7). However, comparisons at these frequencies suggest that theta networks are less specific to recovery of consciousness and states of improved cognitive function, whereas beta networks are less sensitive to recovery of patients in general (Supp. Fig. 7). The functional roles of theta and beta networks may differ from that of alpha in the context of neurological recovery of awareness. Rather than signifying cognitive functional level, theta networks may represent an early prerequisite to orientation and cognitive control^78^, whereas beta networks may represent more transient and functionally specific cognitive states involving frontal regions^79^.

### Limitations and future directions

Coherence-based signatures of network dynamics have potential clinical applications in the diagnosis, prognosis and treatment of patients with DoC^31^. Applied to alpha signals, these measures are sensitive to recovery of consciousness and are potential biomarkers of cognitive function during states of confusion or cognitive motor dissociation. Although previous studies have applied global coherence to higher-density recordings such as 64-channel EEG and electrocorticography^21,23,24^, we successfully derived consistent resting-state measures of brain-wide coherence in low-density 19-channel clinical recordings. Task-relevant effects, in contrast, may require higher-density channel sets as in Curley et al^26^ to achieve greater sensitivity by detecting task-related network interactions at finer scales within spatially confined parietal or occipital regions.

Several limitations to this study arise from the challenges inherent in combining datasets collected by different designs and procedures. Notably, site A’s experiment was initially designed as a study following DoC patients between the acute and chronic settings, whereas site B’s experiment was conducted entirely in the chronic setting. Three CS patients with CRS-R >= 22 and no R-CS patients were recorded in site A’s acute setting; the rest were recorded in the chronic setting. Due to these sample sizes, the distribution of subjects at a given LoC across settings may be a confounder. While data processing procedures were identical between the datasets, other differences between data collection at the two sites, such as inconsistent neuropsychological assessment, original EEG channel sizes, and data segmentation procedures, must be acknowledged. The extent of this limitation is somewhat mitigated by the fact that features we measured were consistent in healthy controls between the two sites.

Alpha network function may be relevant to the development of therapeutic interventions to promote cognitive recovery in patients with DoC. Cortical networks identified by global coherence methods are conceivable targets for next-generation phase-locked brain stimulation, which may be able to reorganize alpha networks by innervating particular regions of cortex at phase offsets determined to be linked to conscious function^80,81^. Subcortically, stimulation of the central thalamus has been shown to increase thalamocortical alpha coherence and restore consciousness in anesthetized macaques^63^, and enhance behavioral responsiveness in a chronic MCS patient^82^, but the extent to which such findings can be applied to cognitive recovery in DoC is unclear. To what degree healthy network interactions require regeneration of specific corticocortical or thalamocortical pathways remains unclear. More detailed electrophysiological and imaging studies are required to decipher the precise layout of relevant cortical and thalamocortical alpha circuitry in the context of DoC.

This study demonstrates the utility of global coherence methods applied to the diagnosis of DoC and establishes a novel network signature of cognitive recovery based on alpha coherence. Global coherence measures, as a natural extension of multitaper spectral analysis, may be used to aid clinical interpretation of EEG generally, given the mechanistic basis of coherence for rhythms such as the alpha. Alpha networks may indicate consciousness-supporting neural circuitry, warranting further study as EEG-based clinical biomarkers.

## Data Availability

The cross-spectral resampling, global coherence, and subspace analytical techniques used in this study can be found in the gcoh+: Global Coherence Toolbox Plus GitHub repository (https://github.com/dvwz/gcoh_plus). Data are available upon request from the corresponding author.

## Acknowledgements

We thank Patrick Purdon for feedback and input regarding the methods and Samuel Snider for helpful comments on the analysis.

## Author Contributions

List of author initials: DWZ, MMC, WHC, CAS, CC, ESR, YGB, JDV, NDS, ENB, BLE MMC, YGB, JDV, NDS, BLE conceived and planned the experiments.

MMC, WHC, CAS, CC, ESR, YGB, NDS, BLE carried out the experiments.

DWZ, MMC, WHC, CAS, CC preprocessed the data.

DWZ, CAS, CC, JDV wrote the preprocessing and analysis software.

DWZ, MMC, WHC, JDV, NDS, ENB, BLE performed the data, statistical, and computational analyses.

DWZ, BLE drafted the manuscript.

DWZ, MMC, WHC, CAS, CC, ESR, YGB, JDV, NDS, ENB, BLE reviewed and edited the manuscript.

DWZ, ENB, BLE funded the project.

## Funding

This study was supported by the NIH T32 Neurobiological Engineering Training Program (T32EB019940, DWZ), NIH Director’s Office ([DP2HD101400, BLE](https://projectreporter.nih.gov/project_info_description.cfm?aid=9781894&cr=1&cs=ASC&csb=default&ddparam=&ddsub=&ddvalue=&icde=46507492&pball=)), James S. McDonnell Foundation (BLE), Tiny Blue Dot Foundation (BLE), and Chen Institute MGH Research Scholars Award (BLE).

## Disclosures

The authors declare no competing interests. The views expressed are purely those of the authors and may not in any circumstances be regarded as stating an official position of the European Research Council Executive Agency (ERCEA) and the European Commission.

## Supplementary Material

**Supplementary Table 1.** Sedative, anxiolytic and analgesic medications administered before and during EEG at study site 1.

| Site + ID | Medications Administered Before EEG | Medications Administered During EEG |
| --- | --- | --- |
| A1 | None | None |
| A2 | N/A | N/A |
| A3 | morphine 2 mg IV | None |
| A4 | propofol 100 mg/hr IV gtt | propofol 100 mg/hr IV gtt + propofol 10 mg IV bolus before motor imagery |
| A5 | None | None |
| A6 | oxycodone 5 mg PGT | quetiapine 25 mg PGT |
| A7 | hydromorphone 0.5 mg IV | None |
| A8 | quetiapine 12.5 mg PGT | None |
| A9 | propofol 300 mg/hr IV gtt | propofol 300 mg/hr IV gtt + hydromorphone 0.5 mg IV |
| A10 | None | None |
| A11 | None | None |
| A12 | lorazepam 1 mg IV + haloperidol 5 mg IV | None |
| A13 | None | None |
| A14 | None | None |
| A15 | propofol 300 mg/hr IV gtt + fentanyl 50 mcg IV bolus | propofol 300 mg/hr IV gtt |
| A16 | N/A | N/A |
| A17 | N/A | N/A |
| A18 | N/A | N/A |
| A19 | N/A | N/A |

**Supplementary Figure 1.**
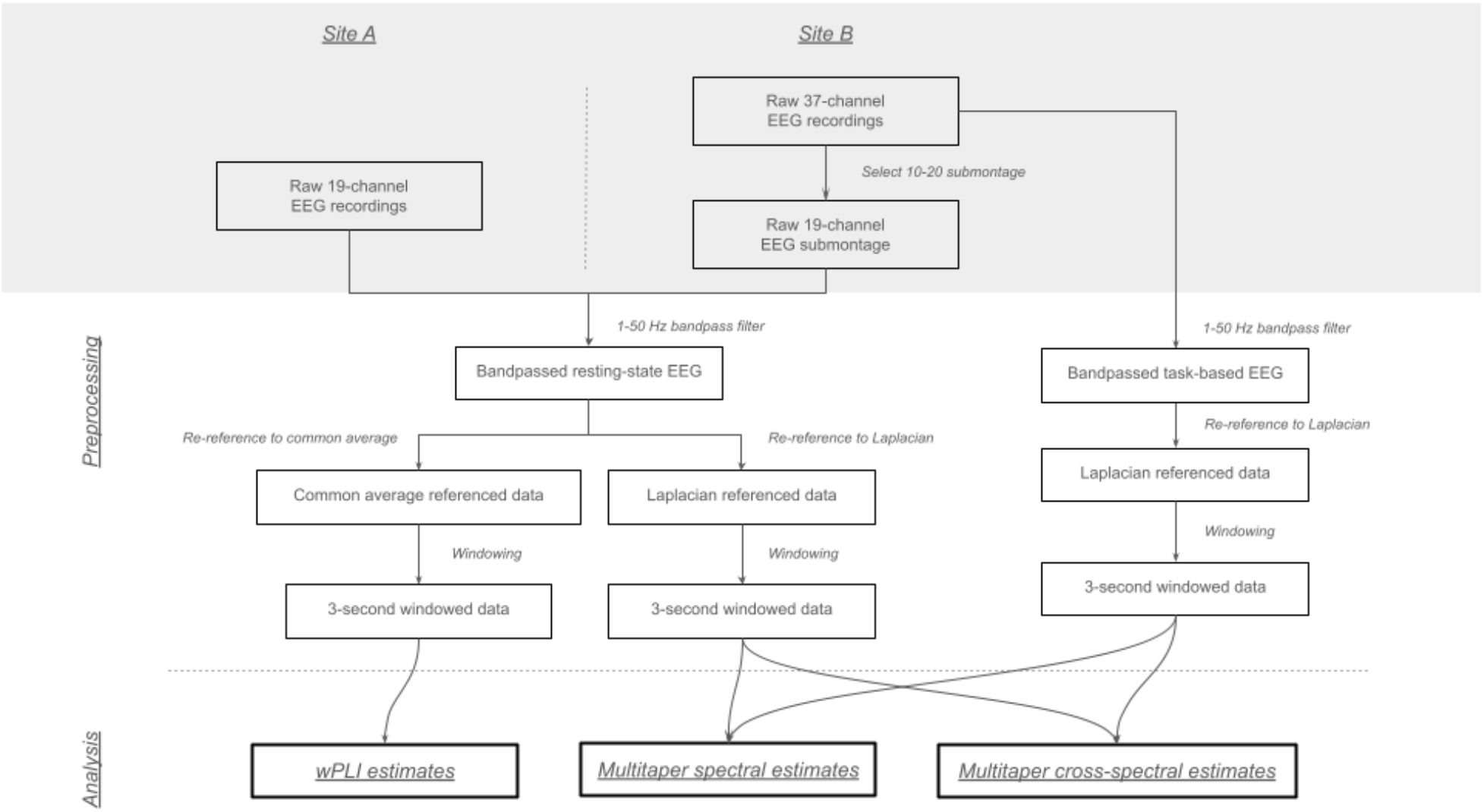
Pre-processing and analysis steps applied to recordings at both study sites.

**Supplementary Figure 2.**
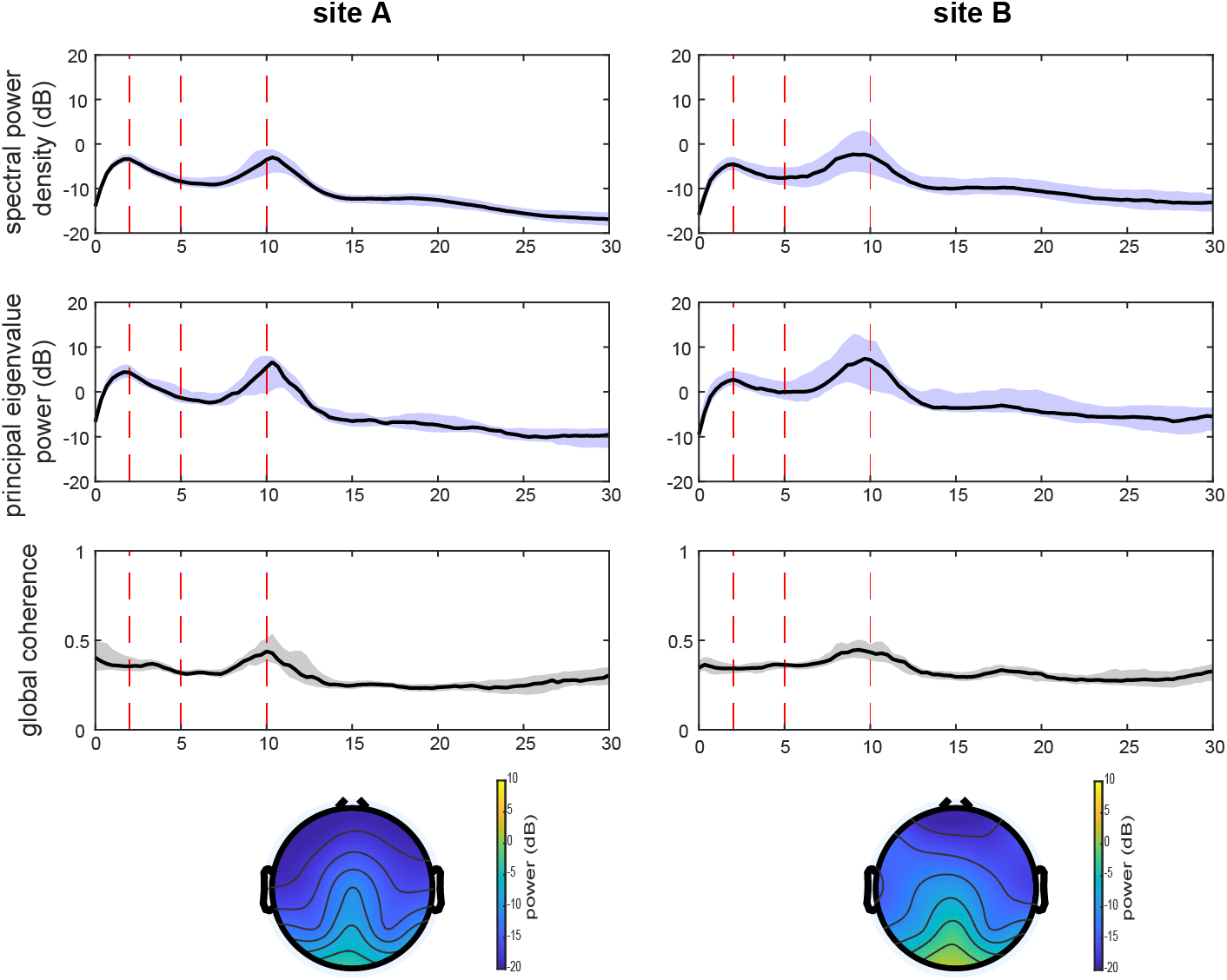
Group-level frequency-domain and topographical plots of spectral power, principal eigenvalue, and global coherence in healthy controls at each experimental site.

**Supplementary Figure 3.**
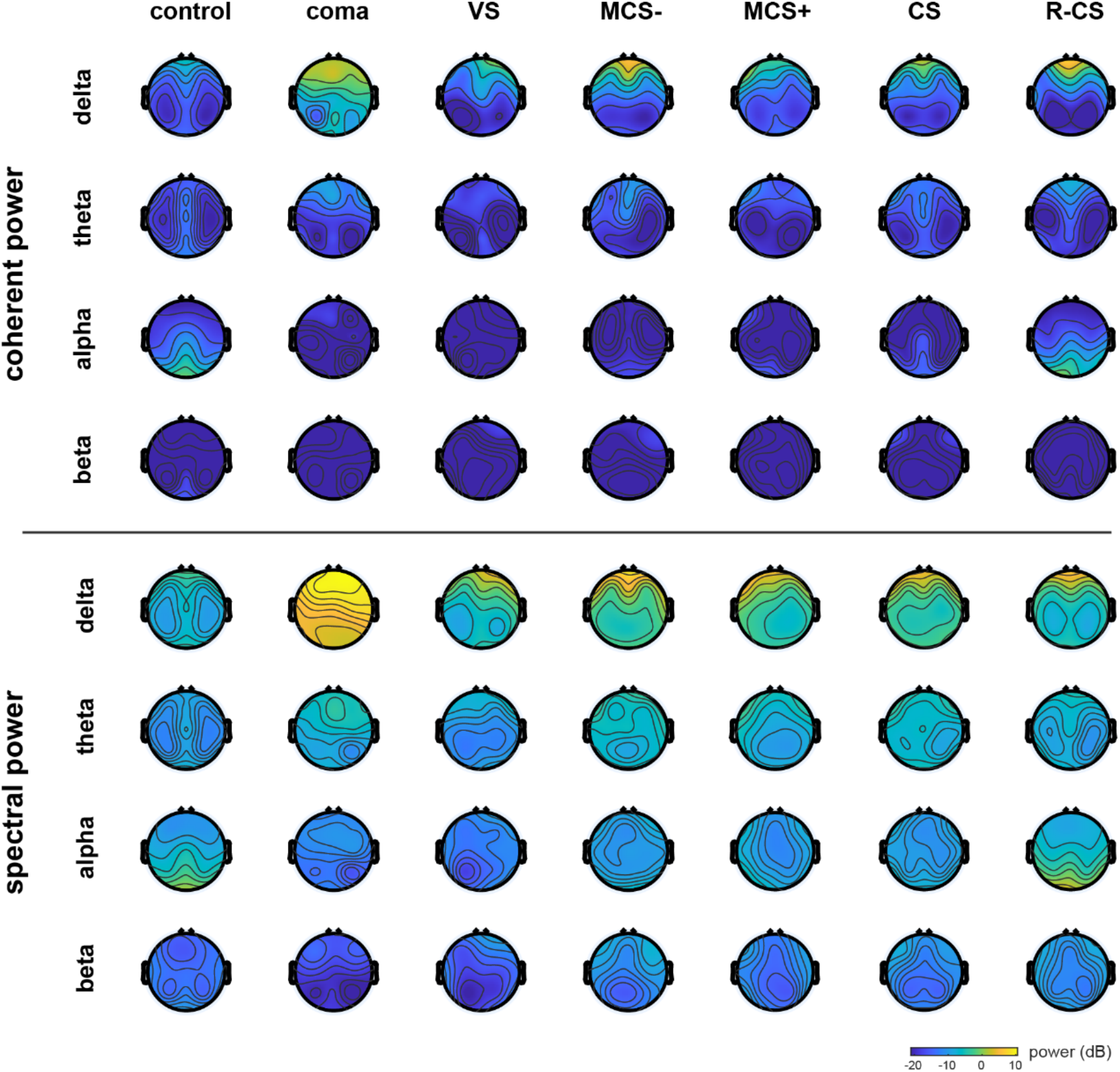
Group-level topographical representations of frequency-dependent spatial components of spectral and coherent spectral power. Representative frequencies were chosen at 2, 6, 10, and 18 Hz representing center frequencies of delta, theta, alpha, and beta bands, respectively.

**Supplementary Figure 4.**
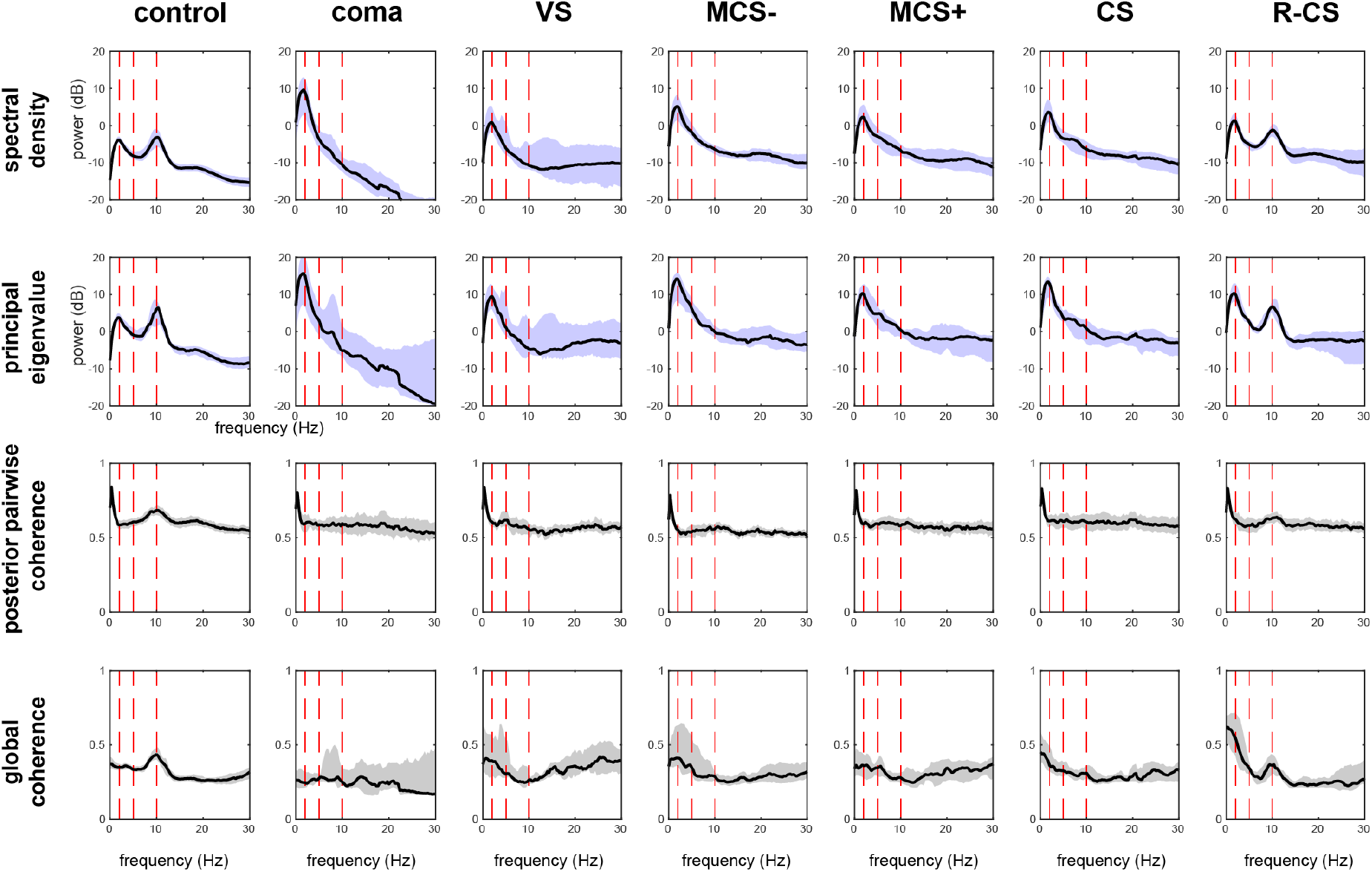
Group-level frequency-domain plots of spectral, cross-spectral, pairwise coherence, and global coherence estimates. Red vertical lines denote the 2, 5, and 10 Hz frequency positions. Posterior pairwise coherence was computed from the mean among the posterior O1-Pz and O2-Pz pairs.

**Supplementary Figure 5.**
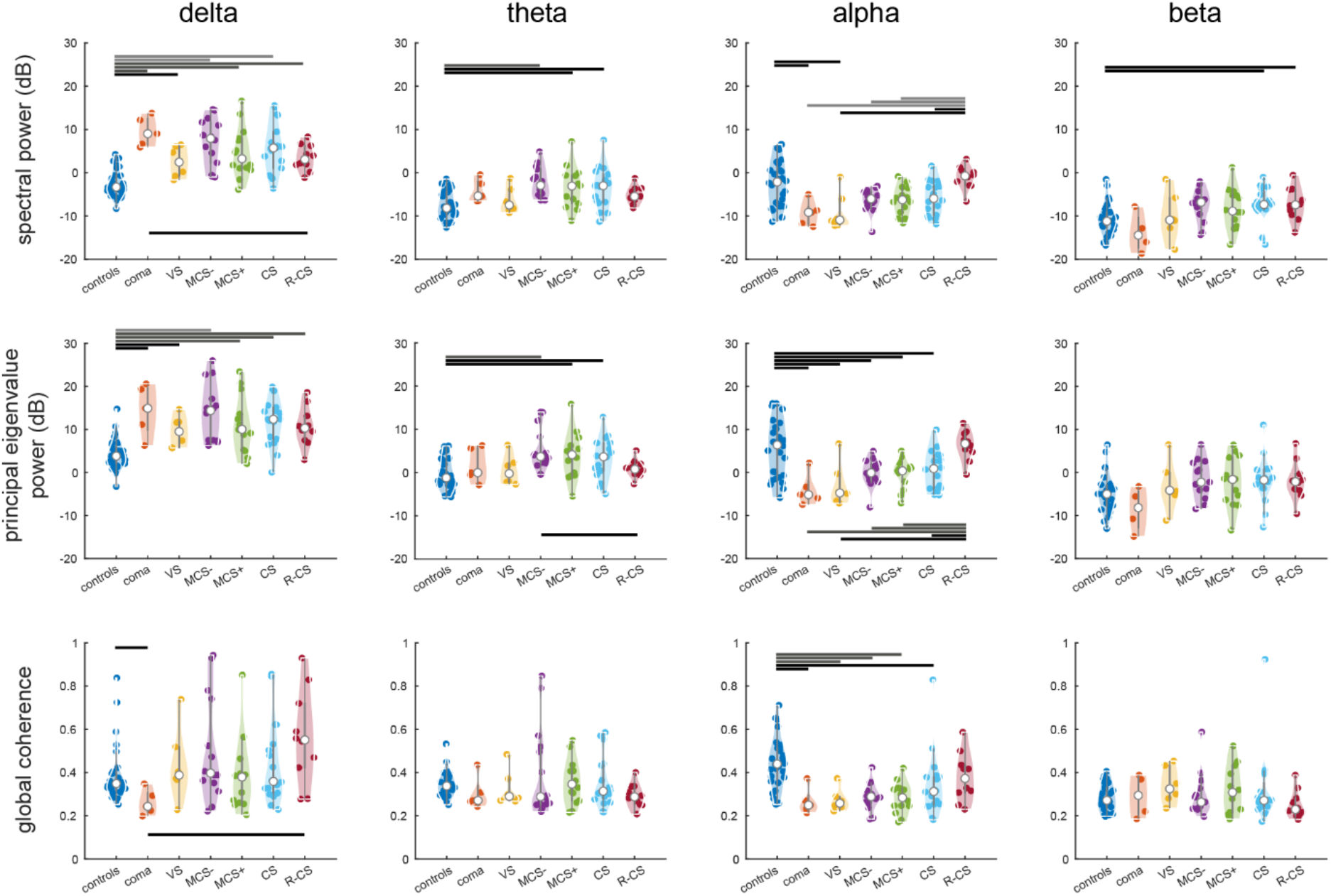
Subject-level spectral, cross-spectral, and global coherence estimates at canonical frequency bands. Each violin plot corresponds to a group-level feature distribution of a given frequency. Dots within each violin plot represent mean estimates for a single subject. Upper horizontal line groups in each subplot denote significant differences between control group distributions and patient group distributions, and lower horizontal line groups represent significant differences between R-CS and DoC group distributions. All hypothesis tests were conducted with two-sided Mann-Whitney U tests, corrected for multiple comparisons using the Benjamini-Hochberg procedure across all frequencies, feature types, and group pairs. Black lines denote significance at a p < 0.05 level, dark gray at a p < 0.01 level, and light gray at a p < 0.001 level, respectively. Definitions of delta, theta, alpha, and beta are identical to those in Supp. Fig. 3.

**Supplementary Figure 6.**
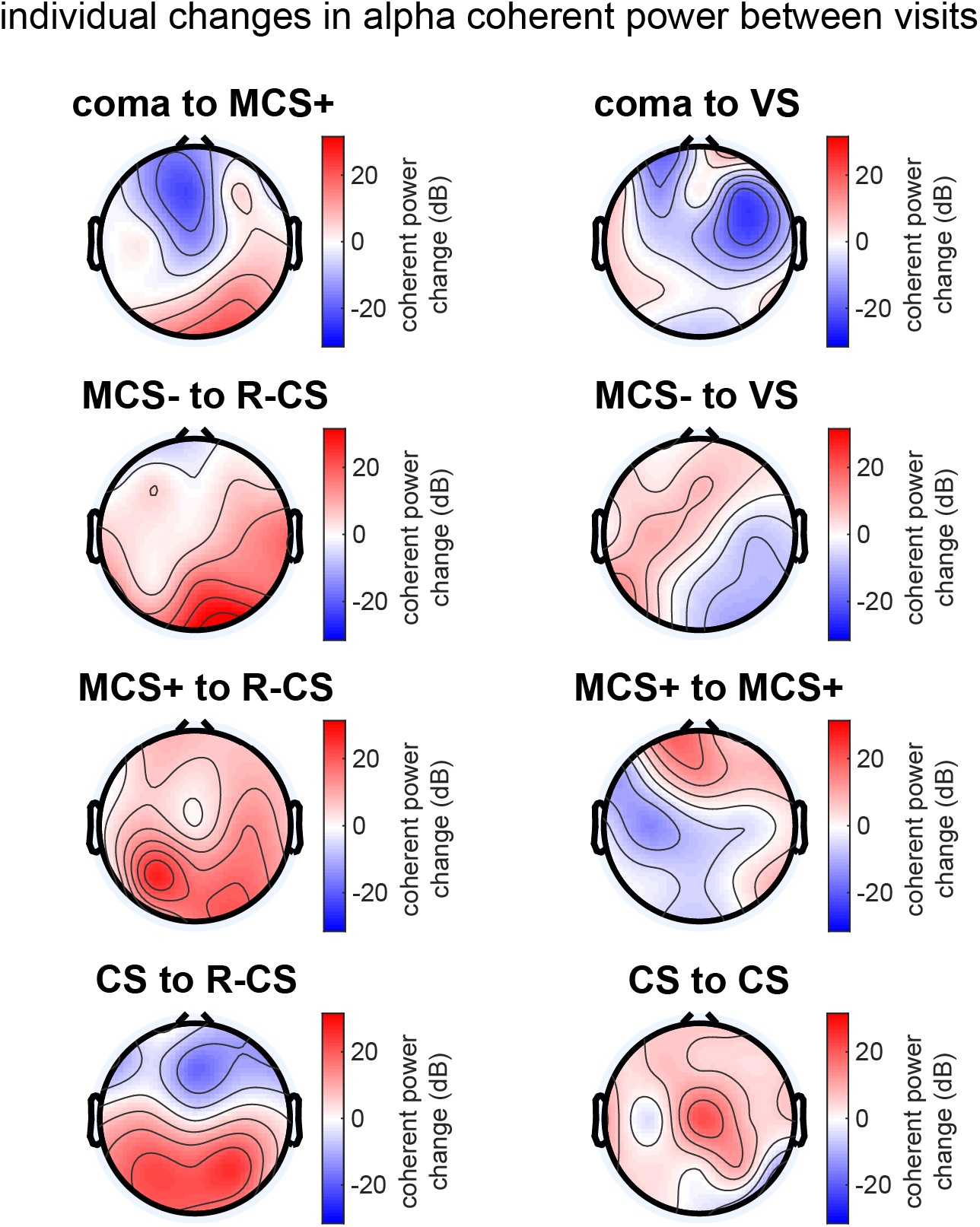
Changes in 10-Hz posterior coherent power between visits among selected individuals. Subjects recover coherent alpha power in posterior channels upon improvement across various levels of consciousness (left). Many subjects who do not improve from DoC to R-CS (right) show lesser degrees of coherent power increase in posterior areas.

**Supplementary Figure 7.**
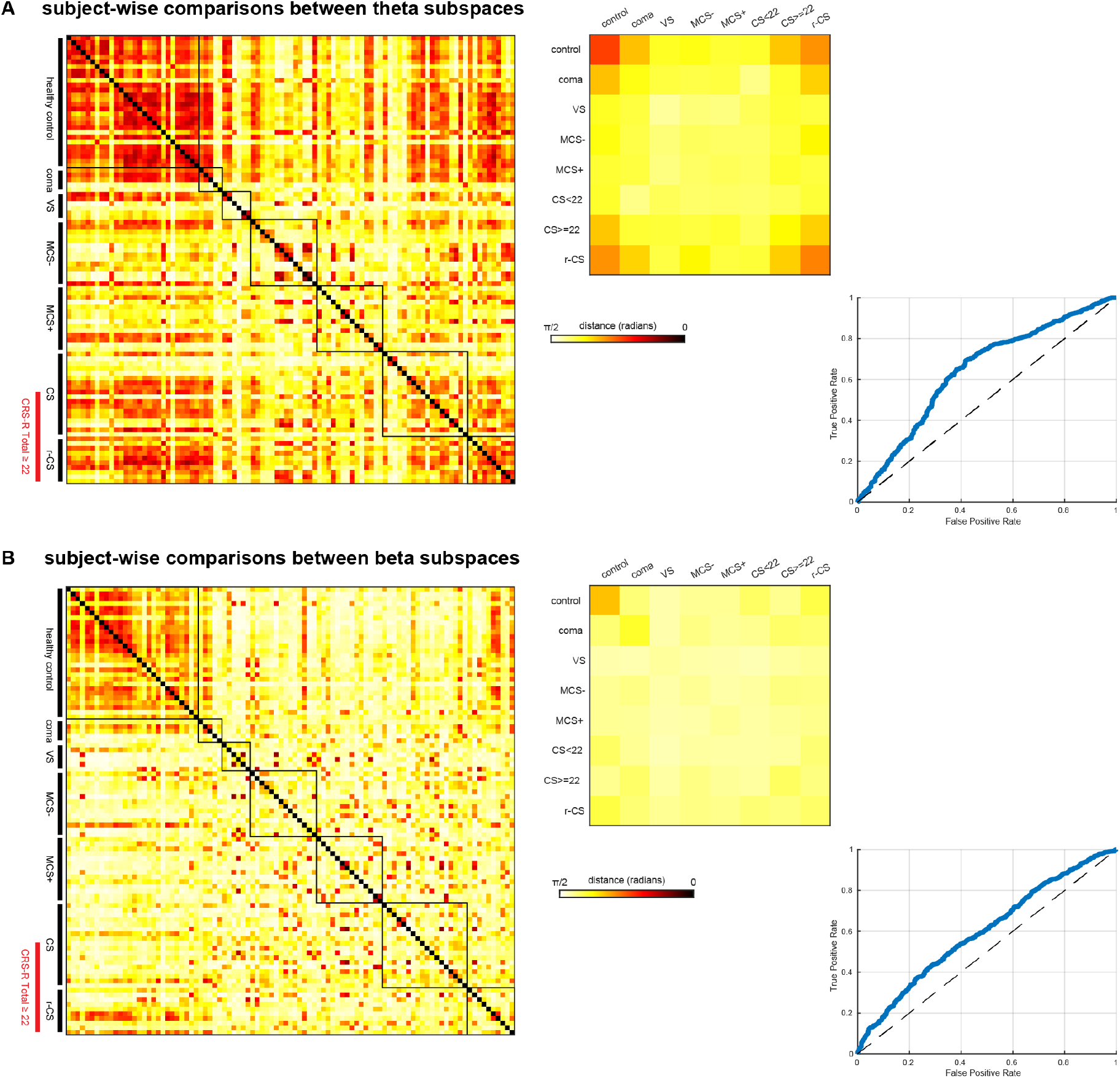
Subject-wise comparisons between subspaces at theta and beta frequencies. Like the alpha, theta and beta rhythms also form coherence-based networks across the brain. However, coherent subspaces at theta are less specific in revealing similarity between conscious subjects, whereas subspaces at beta are less sensitive to similarity between controls and R-CS subjects.

**Supplementary Table 2.** Summary of network-weighted alpha task suppression results in 37-channel EEG recordings of DoC patients and healthy controls. Level of Consciousness (LoC) marked with * represent insufficient testing to ascertain recovery from confusional state (CS). LoC marked with ** determined by clinical emergence from CS. Healthy controls are listed as subjects C1 through C15.

| Subjects | Tasks blocks with alpha suppression (37 ch) | Total task blocks tested | Paradigms with alpha suppression | Total paradigms tested | EEG CF Positive in Curley 2018 | Total CRS-R Score at EEG | LoC |
| --- | --- | --- | --- | --- | --- | --- | --- |
| B21 | 4 | 5 | 2 | 2 | Y | 23 | R-CS** |
| B22 | 1 | 5 | 1 | 3 | N | 8 | MCS- |
| B22 | 2 | 15 | 2 | 3 | Y | 7 | VS |
| B23 | 0 | 5 | 0 | 2 | - | 21 | MCS+ |
| B24 | 2 | 11 | 2 | 3 | - | 7 | VS |
| B25 | 1 | 12 | 1 | 2 | Y | 17 | CS* |
| B25 | 1 | 10 | 1 | 2 | Y | 14 | MCS+ |
| B26 | 1 | 14 | 1 | 3 | Y | 12 | MCS- |
| B26 | 0 | 17 | 0 | 3 | N | 12 | MCS- |
| B27 | 1 | 11 | 1 | 2 | N | 5 | MCS+ |
| B27 | 1 | 16 | 1 | 3 | Y | 5 | MCS+ |
| B28 | 1 | 13 | 1 | 2 | N | 10 | MCS- |
| B29 | 0 | 6 | 1 | 2 | Y | 11 | MCS- |
| B30 | 1 | 9 | 1 | 3 | Y | 17 | MCS+ |
| B31 | 2 | 12 | 2 | 3 | Y | 22 | CS |
| B32 | 1 | 9 | 1 | 2 | - | 23 | CS |
| C1 | 4 | 11 | 3 | 3 |  |  |  |
| C2 | 7 | 11 | 3 | 3 |  |  |  |
| C3 | 5 | 8 | 3 | 3 |  |  |  |
| C4 | 8 | 12 | 3 | 3 |  |  |  |
| C5 | 7 | 9 | 3 | 3 |  |  |  |
| C6 | 7 | 11 | 2 | 3 |  |  |  |
| C7 | 7 | 10 | 3 | 3 |  |  |  |
| C8 | 1 | 5 | 1 | 3 |  |  |  |
| C9 | 4 | 10 | 2 | 3 |  |  |  |
| C10 | 1 | 5 | 1 | 3 |  |  |  |
| C11 | 2 | 5 | 2 | 3 |  |  |  |
| C12 | 8 | 11 | 3 | 3 |  |  |  |
| C13 | 5 | 5 | 3 | 3 |  |  |  |
| C14 | 5 | 9 | 2 | 3 |  |  |  |
| C15 | 3 | 5 | 2 | 3 |  |  |  |

### Software materials and availability

The cross-spectral resampling, global coherence, and subspace analytical techniques used in this study can be found in the *gcoh+: Global Coherence Toolbox Plus* GitHub repository (https://github.com/dvwz/gcoh_plus). Data are available upon request from the corresponding author.

## Notes

### Funding Statement

This study was supported by the NIH T32 Neurobiological Engineering Training Program (T32EB019940, DWZ), NIH Directors Office (DP2HD101400, BLE), James S. McDonnell Foundation (BLE), Tiny Blue Dot Foundation (BLE), and Chen Institute MGH Research Scholars Award (BLE).

### Author Declarations

IRBs of Massachusetts General Hospital and Weill Cornell Medical Center gave ethical approval for this work.

